# Dominant variants in major spliceosome U4 and U5 small nuclear RNA genes cause neurodevelopmental disorders through splicing disruption

**DOI:** 10.1101/2024.10.07.24314689

**Authors:** Caroline Nava, Benjamin Cogne, Amandine Santini, Elsa Leitão, François Lecoquierre, Yuyang Chen, Sarah L. Stenton, Thomas Besnard, Solveig Heide, Sarah Baer, Abhilasha Jakhar, Sonja Neuser, Boris Keren, Anne Faudet, Sylvie Forlani, Marie Faoucher, Kevin Uguen, Konrad Platzer, Alexandra Afenjar, Jean-Luc Alessandri, Stephanie Andres, Chloé Angelini, Bernard Aral, Benoit Arveiler, Tania Attie-Bitach, Marion Aubert Mucca, Guillaume Banneau, Tahsin Stefan Barakat, Giulia Barcia, Stéphanie Baulac, Claire Beneteau, Fouzia Benkerdou, Virginie Bernard, Stéphane Bézieau, Dominique Bonneau, Marie-Noelle Bonnet-Dupeyron, Simon Boussion, Odile Boute, Elise Brischoux-Boucher, Samantha J. Bryen, Julien Buratti, Tiffany Busa, Almuth Caliebe, Yline Capri, Kévin Cassinari, Roseline Caumes, Camille Cenni, Pascal Chambon, Perrine Charles, John Christodoulou, Cindy Colson, Solène Conrad, Auriane Cospain, Juliette Coursimault, Thomas Courtin, Madeline Couse, Charles Coutton, Isabelle Creveaux, Alissa M. D’Gama, Benjamin Dauriat, Jean-Madeleine de Sainte Agathe, Giulia Del Gobbo, Andree Delahaye-Duriez, Julian Delanne, Anne-Sophie Denommé-Pichon, Anne Dieux-Coeslier, Laura Do Souto Ferreira, Martine Doco-Fenzy, Stephan Drukewitz, Véronique Duboc, Christèle Dubourg, Yannis Duffourd, David Dyment, Salima El Chehadeh, Monique Elmaleh, Laurence Faivre, Samuel Fennelly, Hanna Fischer, Mélanie Fradin, Camille Galludec Vaillant, Benjamin Ganne, Jamal Ghoumid, Himanshu Goel, Zeynep Gokce-Samar, Alice Goldenberg, Romain Gonfreville Robert, Louise Goujon, Victoria Granier, Mathilde Gras, John M. Greally, Bianca Greiten, Paul Gueguen, Anne-Marie Guerrot, Saurav Guha, Anne Guimier, Tobias Haack, Hamza HadjAbdallah, Yosra Halleb, Radu Harbuz, Madeleine Harris, Julia Hentschel, Bénédicte Héron, Marc-Phillip Hitz, A. Micheil Innes, Vincent Jadas, Louis Januel, Nolwenn Jean-Marçais, Vaidehi Jobanputra, Florence Jobic, Ludmila Jornea, Céline Jost, Sophie Julia, Frank J. Kaiser, Daniel Kaschta, Sabine Kaya, Petra Ketteler, Bochra Khadija, Fabian Kilpert, Cordula Knopp, Florian Kraft, Ilona Krey, Marilyn Lackmy, Fanny Laffargue, Laetitia Lambert, Ryan Lamont, Vincent Laugel, Steven Laurie, Julie L. Lauzon, Louis Lebreton, Marine Lebrun, Marine Legendre, Eric Leguern, Daphné Lehalle, Elodie Lejeune, Gaetan Lesca, Marion Lesieur-Sebellin, Jonathan Levy, Agnès Linglart, Stanislas Lyonnet, Kevin Lüthy, Alan S. Ma, Corinne Mach, Jean-Louis Mandel, Lamisse Mansour-Hendili, Julien Marcadier, Victor Marin, Henri Margot, Valentine Marquet, Angèle May, Johannes A. Mayr, Vincent Michaud, Caroline Michot, Gwenael Nadeau, Sophie Naudion, Laetitia Nguyen, Mathilde Nizon, Frédérique Nowak, Sylvie Odent, Valerie Olin, Ikeoluwa A. Osei-Owusu, Matthew Osmond, Katrin Õunap, Laurent Pasquier, Sandrine Passemard, Olivier Patat, Marine Pensec, Laurence Perrin-Sabourin, Florence Petit, Christophe Philippe, Marc Planes, Annapurna Poduri, Céline Poirsier, Antoine Pouzet, Bradley Prince, Clément Prouteau, Aurora Pujol, Caroline Racine, Mélanie Rama, Francis Ramond, Kara Ranguin, Margaux Raway, Mathilde Renaud, Nicole Revencu, Anne-Claire Richard, Lucile Riera-Navarro, Rocio Rius, Diana Rodriguez, Agustí Rodriguez-Palmero, Sophie Rondeau, Annika Roser-Unruh, Christelle Rougeot Jung, Hana Safraou, Véronique Satre, Pascale Saugier-Veber, Clément Sauvestre, Elise Schaefer, Wanqing Shao, Ina Schanze, Jan-Ulrich Schlump, Agatha Schlüter Martin, Caroline Schluth-Bolard, Christopher Schröder, Monisha Sebastin, Sabine Sigaudy, Malte Spielmann, Marta Spodenkiewicz, Laura St Clair, Julie Steffann, Radka Stoeva, Harald Surowy, Mark A. Tarnopolsky, Calina Todosi, Annick Toutain, Frédéric Tran Mau-Them, Astrid Unterlauft, Julien Van-Gils, Clémence Vanlerberghe, Gabriella Vera, André Verdel, Alain Verloes, Yoann Vial, Cédric Vignal, Marie Vincent, Catherine Vincent-Delorme, Antonio Vitobello, Sacha Weber, Marjolaine Willems, Khaoula Zaafrane-Khachnaoui, Pia Zacher, Lena Zeltner, Alban Ziegler, Wojciech P. Galej, Hélène Dollfus, Christel Thauvin, Kym M. Boycott, Pierre Marijon, Alban Lermine, Valérie Malan, Marlène Rio, Alma Kuechler, Bertrand Isidor, Séverine Drunat, Thomas Smol, Nicolas Chatron, Amélie Piton, Gael Nicolas, Matias Wagner, Rami Abou Jamra, Delphine Héron, Cyril Mignot, Pierre Blanc, Anne O’Donnell-Luria, Nicola Whiffin, Camille Charbonnier, Clément Charenton, Julien Thevenon, Christel Depienne

## Abstract

Variants in *RNU4-2*, encoding the small nuclear RNA (snRNA) U4, were recently identified as a major cause of neurodevelopmental disorders (ReNU syndrome). Here, we investigated *de novo* variants in 50 snRNAs in a French cohort of 23,649 individuals with rare disorders and collected data of additional patients through an international collaboration. Altogether, we identified 133 probands with pathogenic or likely pathogenic variants in *RNU4-2* and 15 individuals with *de novo* and/or recurrent variants in constrained regions of *RNU5B-1*, one of five genes encoding U5. These variants cluster in evolutionarily conserved regions of U4 and U5 essential for splicing. *RNU4-2* variants affecting stem III are associated with milder phenotypes than those in the T-loop (quasi-pseudoknot). Phaseable variants associated with severe phenotypes occurred on the maternal allele. Individuals with *RNU4-2* variants show specific defects in alternative 5’ splice site usage, correlating with variant location and clinical severity. Additionally, we report an episignature associated with severe ReNU syndrome. This study further highlights the importance of *de novo* variants in snRNAs and establishes *RNU5B-1* as a new neurodevelopmental disorder gene.

## Introduction

The splicing of pre-mRNA into mature mRNA in eukaryotic cells consists of excising introns and ligating exons. The two transesterification reactions necessary for this process are carried out by a large ribonucleoprotein (RNP) complex called the spliceosome^1,2^. This complex is composed of five uridyl-rich small nuclear RNAs (snRNAs) that are essential for spliceosome assembly and function and differ according to the type of excised intron. The major spliceosome processes the majority (>99%) of introns containing GU-AG splice sites (U2-type) and is composed of snRNAs U1, U2, U4, U5 and U6^3^. Each snRNA has unique sequence motifs and secondary structures that allow it to interact precisely with its target sites. U1 and U2 respectively bind to the 5’ splice sites and branch points, while U4, U5, and U6 form the tri-snRNP complex that is recruited to assemble a precatalytic spliceosome complex. U6 is initially maintained in an inactive conformation by pairing with U4. Dissociation of the U4/U6 interaction allows U6 to interact with U2 and form the catalytic site^4^. U5 is responsible for aligning the exons for ligation by binding to the 5’ and 3’ splice sites, ensuring accurate exon joining^5^.

Spliceosomal snRNAs are ubiquitously expressed and encoded by distinct single-exon genes transcribed by polymerase II and/or polymerase III^6^. Genes encoding snRNAs U1, U2, U4, U5 and U6 in human genomes are present in multiple copies, some of which are functional and others are pseudogenes^7,8^. After transcription, snRNAs undergo extensive processing steps essential for their stability and function. These include 5’-capping, 3’-end processing, nuclear export, binding to the Sm protein/SMN complex via their conserved Sm site to form the core snRNP structure, nuclear re-import, and nucleotide modifications (2’-O-methylation, and pseudouridylation) guided by small Cajal body-specific RNAs (scaRNAs)^9-12^.

A recent landmark discovery has implicated *de novo* variants in *RNU4-2*, one of two functional genes encoding U4, as a major cause of a neurodevelopmental disorder (NDD) called ReNU syndrome (OMIM#620851)^13,14^. Genome sequencing is necessary to detect these variants, which are typically not captured by current exome sequencing. The identification of variants in *RNU4-2* was facilitated by the high recurrence of a single base insertion (n.64_65insT), which represented 78% of all pathogenic variants identified in patients. This variant is statistically enriched in the Genomics England (GEL)^15^ NDD cohort while absent from gnomAD^16^ and highly depleted in UK Biobank^17^. The analysis of 28 other brain-expressed snRNA genes did not reveal any enrichment of *de novo* variants similar to *RNU4-2*, although 14 regions of 13 genes appear more evolutionary constrained^13^. These results raise the question of whether variants in other snRNA genes may underlie other rare diseases, and how to accurately classify variants in these genes.

In this study, we investigated snRNA genes in a French cohort of 23,649 patients with various rare disorders and collected variants in additional cohorts via international collaborations. Using these data, we implicated one further snRNA in NDDs and more comprehensively defined ReNU syndrome. Additionally, we investigated *RNU4*-2 splicing profiles by RNA sequencing and performed DNA methylation studies, identifying a ReNU syndrome-associated episignature.

## Results

### Analysis of *RNU4-2* variants in multiple cohorts of patients with rare diseases

We investigated *de novo* variants in *RNU4-2* (NR_003137.2) and/or rare variants (<10 alleles in gnomADv4.1.0) located in the 18-bp critical region defined by Chen *et al*.^13^ in the Plan France Médecine Génomique 2025 (PFMG) cohort comprising 23,649 patients with rare disorders (15,073 with NDD). This analysis revealed 73 patients with *de novo RNU4-2* variants. Among the patients for whom parental analysis was not possible, four had variants previously reported as *de novo* in another unrelated individual, and one patient had a single nucleotide deletion (n.76del) within the critical region.

In parallel, we collected data of 60 patients with *RNU4-2* variants identified through genome sequencing data reanalysis (22 patients) or targeted sequencing (38 patients including one monozygotic twin pair). Variants occurred *de novo* in 44/45 cases for whom both parents were available. One patient had a variant (n.72_73del) inherited from an affected father, which had occurred *de novo* in another unrelated patient.

Altogether, 138 individuals had 22 distinct *RNU4-2* variants. Ninety-five patients (43 males, 52 females including the twin; 69%) had the recurrent n.64_65insT insertion. Seven other variants were recurrent: n.76C>T (*n*=9), n.66A>G (*n*=5), n.67A>G (*n*=5), n.65A>G (*n*=3), n.77_78insT (*n*=3), n.70T>C and n.72_73del (2 patients each). Fourteen *de novo* variants were identified in single patients. All but three variants clustered in the 18-bp critical region spanning nucleotides 62 to 79 (chr12(hg38):120,291,825-120,291,842)^13^. This region overlaps four distinct domains in the U4/U6 structure^4^ (Fig. 1): stem I (U4 n.62), T-loop/quasi-pseudoknot (n.63-67), RBM42 interaction region (n.68-70) and stem III (n.72-79). The remaining variants were located in the 5’ stem loop between stem I and stem II (n.45_46insT) or in the 3’ stem loop (n.92C>G, n.111C>T). Eighteen variants were absent from gnomADv4.1.0, AllofUs, and TOPMed_freeze10 while three were present at very low frequency (n.66A>G, n.76C>T once each; n.76del: 3 times; Supplementary Fig. 1A; Supplementary Table 1). Two variants (both located outside of the 18-bp critical region) had higher occurrences: n.92C>G and n.111C>T, 16 and 82 times respectively in gnomADv4.1.0, AllofUs and TOPMed combined. We classified 18 variants (in 134 individuals; 133 probands) as pathogenic (P) or likely pathogenic (LP) using ACMG/AMP criteria^18,19^ and four variants (n.45_46insT, n.92C>G, n.111C>T, n.76del) as of uncertain significance (methods).

**Figure 1.**
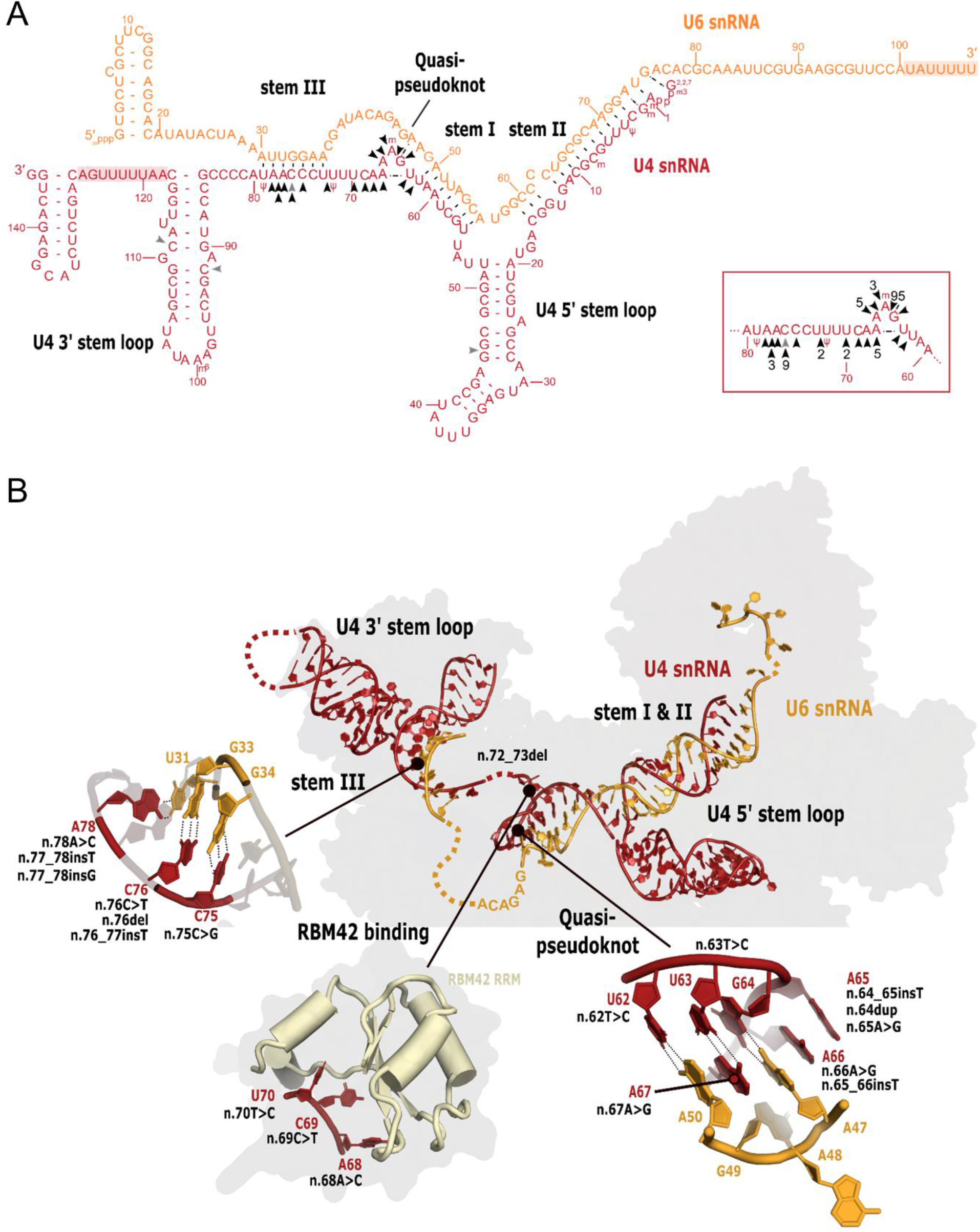
Overview of *RNU4-2* variants identified in this study. **(A)** Two-dimensional predicted structure of the interaction between U4 (red) and U6 (orange) small nuclear RNAs showing distinct domains. Arrows indicate variants from this study (pathogenic and likely pathogenic in black, variants of uncertain significance (VUS) in grey). Numbers in black in the zoom-in represent the number of patients with the variant, for nucleotide changes appearing more than once. The four nucleotide differences between *RNU4-2* and *RNU4-1* are shown using IUPAC codes. Red and orange numbers refer to the numbering of nucleotides from each snRNA. Ψ: pseudouridine, m: 2’-O-methyl residues; m6: N6-methyladenosine; ^2,2,7^m3Gppp: 2,2,7-trimethylguanosine cap; mpppG: 5’ guanosine triphosphate cap with gamma-monomethyl phosphate. Shaded regions: Lsm/Sm protein binding sites. **(B)** Organisation of the U4-U6 duplex at the tri snRNP stage (PDB 6QW6) and close-up views of stem III, RBM42 binding and quasipseudoknot regions. Interactions stabilizing these structures as well as mutations potentially affecting their stability are represented.

We assessed CADD PHRED scores, nucleotide conservation, and *in silico* structure predictions for the 18 LP/P variants and compared these features with variants in *RNU4-2* present in gnomADv4.1.0. While no differences were found in CADD scores or vertebrate conservation (verPhyloP), LP/P variants had a greater predicted effect on U4/U6 interaction compared to variants observed ≥ 10 times in gnomAD (Supplementary Fig. 2; Supplementary Table 2). However, due to the large overlap, these predictions cannot be used to predict variant pathogenicity.

### *De novo* analysis in snRNA genes reveals variants in brain-expressed U5 genes

We next analysed *de novo* variants in 49 additional genes encoding snRNAs (Supplementary Table 3) in the PFMG cohort. This analysis revealed 12 rare *de novo* and/or recurrent alterations in eight genes in 15 unrelated patients. Notably, eight variants (in 11 patients) were located in genes encoding U5 snRNAs (two in *RNU5A-1* (NR_002756.2), four in *RNU5B-1* (NR_002757.3), one in *RNU5E-1* (NR_002754.2) and one in *RNU5F-1* (NR_002753.5)). Three variants (*RNU5A-1* n.40_41insA, *RNU5B-1* n.39C>G and n.44A>G) were recurrent and identified in two individuals each. In seven patients, the variant was located in the highly conserved 5’ loop I (Fig. 2), which is depleted in variants in gnomADv4.1.0 (Supplementary Fig. 1B) and in the UK Biobank^13^. Four *de novo* variants were located in other parts of U5, the internal loops 1 and 2 (*RNU5B-1* n.24G>C, and n.74T>C), the variable 3’ stem loop II (*RNU5F-1* n.115C>A), or the Sm site (*RNU5E-1* n.90_91insA). The individual with *RNU5F-1* n.115C>A had a *de novo* heterozygous frameshift variant in *KDM5B*. All other patients remained negative after genomic variant analysis, suggesting the variants in U5 encoding genes as possible underlying genetic causes.

**Figure 2.**
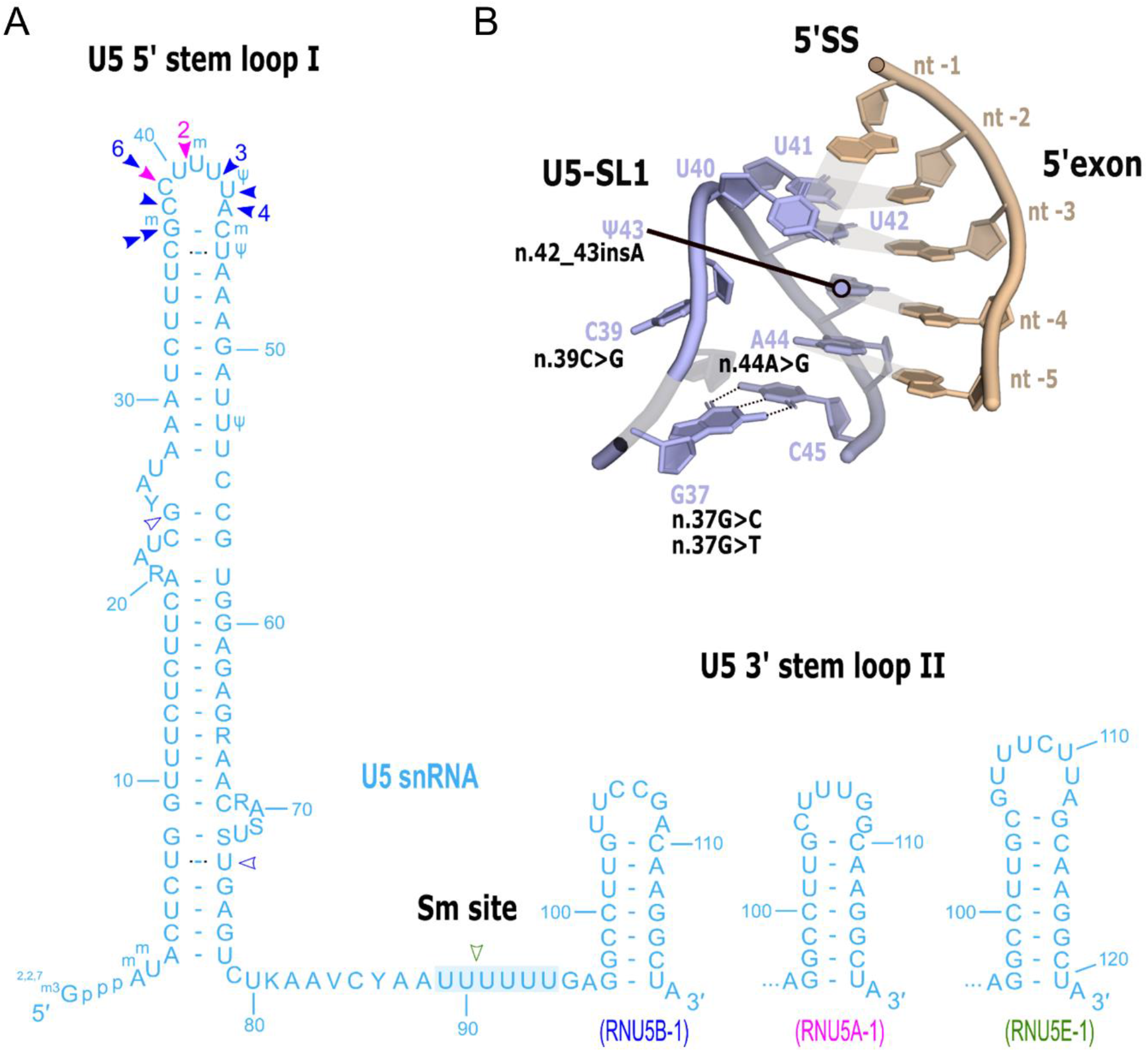
Overview of *RNU5A-1, RNU5B-1* and *RNU5E-1* variants identified in this study. Two-dimensional predicted structure of U5 (blue) small nuclear RNA showing distinct domains. Arrows indicate variants from this study (pink: *RNU5A-1*, dark blue: *RNU5B-1*, green: *RNU5E-1*; pathogenic and likely pathogenic with filled color, variants of uncertain significance (VUS) filled in white). Numbers near the arrows represent the number of patients with the variant, for nucleotide changes appearing more than once. Nucleotide differences between *RNU5A-1, RNU5B-1* and *RNU5E-1* are shown using IUPAC codes, except for the highly variable 3’ stem loop II, for which we show separate loop. Blue numbers refer to the numbering of nucleotides from each snRNA. Ψ: pseudouridine; m: 2’-O-methyl residues; ^2,2,7^m3Gppp: 2,2,7-trimethylguanosine cap. Shaded region: Sm site. (**B**) 5’exon recognition by the U5 stem loop I (SL1) at the B-complex stage (PDB 8Q7N).

In parallel, we collected information on rare *de novo* variants in *RNU5A-1* and *RNU5B-1* in data from the 100,000 Genomes Project (GEL) and NHS GMS cohorts available within Genomics England (Supplementary Table 5). This analysis identified five *de novo* variants in *RNU5B-1* in six NDD probands (n.37G>C; n.37G>T; n.42_43insA twice, n.44A>G; n.64G>A). This was compared to only a single non-NDD individual in GEL / NHS GMS having a *de novo* variant in *RNU5B-1* (n.59G>C; 6/12,724 undiagnosed NDD vs 1/30,058 non-NDD; Fisher’s *p*-value = 0.0036). Additionally, we collected data of five patients with *de novo* variants (n.39C>G: 3 patients; n.42_43insA; n.44A>G) from additional cohorts, including the Broad Centre for Mendelian Genomics (two patients), the BCH Epilepsy Genetics Program, the Australian undiagnosed network, and Care4Rare Canada (one patient each).

Altogether, 13 NDD probands had five distinct *de novo* variants in the 5’ loop I of *RNU5B-1* (Supplementary Table 6). These variants recurrently affected the same nucleotide positions: n.37G>C and n.37G>T, each found in one individual; n.39G>C (five patients); n.42_43insA and n.44A>G (three patients each). This led us to define a critical region in *RNU5B-1* spanning chr15(hg38):65,304,713-64,304,720. Three additional patients analysed in duo in the GEL/NHS GMS cohort and one patient in the PFMG had variants located within this critical region that were absent from the available sequenced parent. In contrast, no individuals with non-NDD phenotypes in GEL/NHS GMS had de novo or duo variants in this region (8/12,724 undiagnosed NDD vs 0/30,058 non-NDD in GEL/NHS GMS; Fisher’s p-value = 6.1×10^−5^). In total, we identified 17 NDD individuals with variants in this critical region of *RNU5B-1*. No other *de novo* variants were identified in *RNU5A-1* in NDD probands. *RNU5A-1* or *RNU5B-1* are the main genes encoding U5 in the brain (Supplementary Fig. 3).

### Most pathogenic *RNU4* and *RNU5* variants occur *de novo* on the maternal allele

We investigated the parental origin of *RNU4-2* variants in available genome data by phasing *de novo* variants and informative SNPs in the flanking regions (methods; Supplementary Fig. 4). We could reliably determine the parental origin of the mutations in 45 trios. The variant was assigned to the maternal allele in 42 cases and to the paternal allele in only three cases. Notably, all 34 n.64_65insT variants were phased to the maternal allele, consistent with observations from Chen *et al*.^13^. Among the variants assigned to the paternal allele, two were classified as likely pathogenic (n.62T>C, n.68A>C) and one (n.92C>G) was of uncertain significance. All three paternally derived variants were SNVs, while this variant type only represented 22% (10/45) of phased variants (Fisher’s *p*-value = 0.0085).

Among the phaseable variants located in the 5’ loop I *RNU5B-1*, five (n.39C>G, n.42_43insA and three n.44A>G) were phased to the maternal allele, while two (n.39C>G. n.37G>C) were on the paternal allele. The two *RNU5B-1 de novo* variants located outside of the conserved 5’ loop I (n.24G>C, n.74T>C) were also phased to the paternal allele. Both n.40_41insA variants in *RNU5A-1* occurred *de novo* on the maternal allele.

### *RNU4-2* variants in the T loop (quasi-pseudoknot) and stem III differ in severity

Clinical data were available for 129 patients with P/LP variants in *RNU4-2* (63 males, 66 females excluding the monozygotic twin who had an identical phenotype to her sister). The median age at study entry was 9 years (range: 4m-45y). All patients had NDD with intellectual disability (ID) of variable severity, ranging from mild (8%), moderate (29%) to severe/profound (63%).

We investigated genotype-phenotype correlations to understand the phenotypic variability of *RNU4-2*-related disorders. We first performed an unsupervised clustering of clinical features, which revealed two separate clusters with different phenotypic severity (Fig. 3A). Most *RNU4-2* variants in stem III (71%, 12/17) were in the mild phenotype cluster, whereas most variants in the T-loop and RBM42 interacting region were in the higher severity cluster (95%, 106/111). The same finding was observed when principal component analysis (PCA) was performed. Variants in stem III and variants in the T-loop separated on the first PC axis, accounting for 12.3% of the variance (Fig. 3B-C; Supplementary Fig. 5). These results suggest that a large part of the phenotypic variability is due to the location of *RNU4-2* variants in different U4 functional domains.

**Figure 3.**
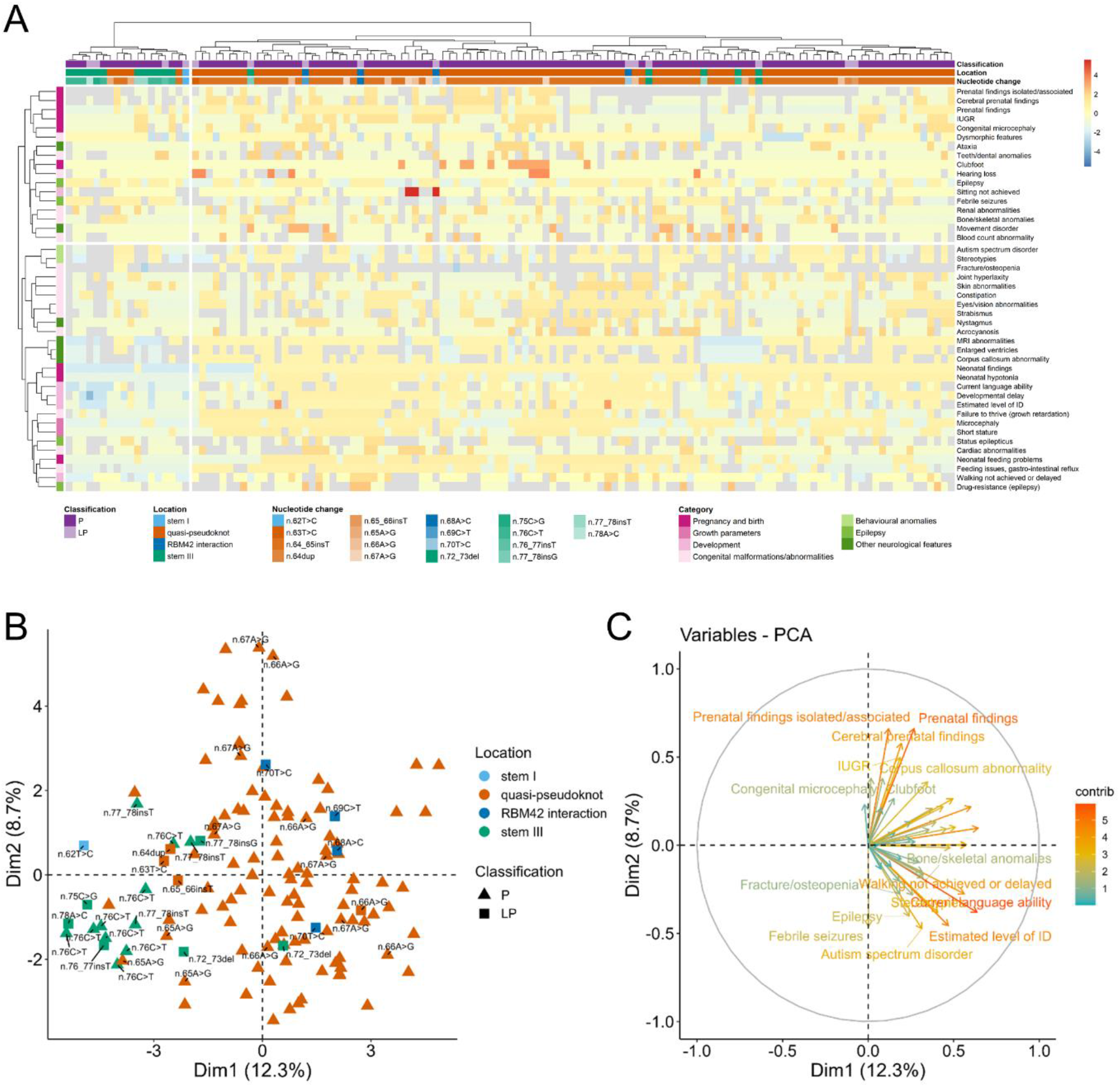
*RNU4-2* variants in T loop and stem III are associated with different phenotype severity. **(A)** Hierarchical clustering of the clinical features (*n*=44, rows) of patients with pathogenic (P) and likely pathogenic (LP) *RNU4-2* variants (*n*=129, columns). Categorical data was converted to 0-1 scale, and values were Z-score scaled for each row. Blue-yellow-red scale depicts Z-scores. Lower values indicate a more favorable phenotype, while higher values represent a more severe phenotype. Missing values are shown in grey. Columns are colored based on the variant classification (purple: P, light purple: LP), its location within the distinct U4:U6 domains (stem I: light blue, quasi-pseudoknot: orange, RBM42 interaction region: blue, stem III: green) and the nucleotide change (color shades related to their position within the respective U4:U6 domain). Rows are colored on the category of the clinical feature (shades of pink and green). **(B)** Principal component analysis (PCA) showing the separation of variants with respect to their location within distinct U4:U6 domains. Labels with the nucleotide change appear for variants other than n.64_65insT. Colored according to the location within the distinct U4:U6 domains: stem I, light blue; quasi-pseudoknot, orange; RBM42 interaction region, blue; stem III, green). Triangles: P variants, squares: LP variants. **(C)** Contributions of the clinical features for the PCA.

Clinical data were available for 91 patients with *RNU4-2* c.64_65insT (Table 1; Supplementary Table 7; Supplementary Fig. 6). Prenatal findings were observed in 50/82 (61%) of cases, predominantly intrauterine growth restriction (IUGR) (28%) and cerebral abnormalities (32%), mainly ventriculomegaly (18%). Of the foetuses with abnormalities, 62% had isolated signs, while 38% exhibited two or more signs, including IUGR, cerebral abnormalities, talipes equinovarus, and renal abnormalities. Neonatal findings were frequent (90%), with hypotonia (69%) and feeding difficulties (57%) being the most common. Microcephaly (head circumference (HC) < 3^rd^ percentile) was present at birth in 26%, and at last examination in 74% (Supplementary Fig. 7). Short stature was noted in 58% of individuals. All patients older than three years (*n*=73) exhibited developmental delay. Most could walk, with a median walking age of 32 months (range 13 months to 12 years), whereas 14% did not reach this milestone. Most patients were non-verbal (60%) or could only speak a few words (35%). The majority had severe/profound ID (77%), with 22% having moderate ID and one patient having mild ID. Behavioural disturbances were common, with 83% displaying autistic features and/or midline stereotypies reminiscent of Rett syndrome in some patients. Epilepsy affected 57%, with an additional 9% experiencing a single seizure. Seizures typically started between 15 months and 8 years (median onset at 33 months) and were usually generalized, rare, fever-sensitive, and responsive to antiepileptic medications. However, five patients were diagnosed with developmental and epileptic encephalopathy; 14 experienced status epilepticus, and seven had drug-resistant epilepsy. Nystagmus and cerebellar ataxia were observed in 36% and 23%, respectively. Brain MRI abnormalities were prevalent (90%), with the most common findings being enlarged ventricles (60%) and a thin corpus callosum (34%). Less common findings included heterotopia (*n*=7), delayed myelination or hypomyelination (*n*=7), and abnormal gyration (*n*=4). Cardiac and genitourinary or renal abnormalities were less common, affecting 19% and 18% of patients respectively, and were generally benign. Skeletal abnormalities, including osteopenia or fractures (*n*=18) and hip dysplasia (*n*=9), were seen in 40% of cases. Dysmorphic features could suggest Pitt-Hopkins syndrome. Strabismus, and drooling were also common. Feeding difficulties affected 69%, failure to thrive 55%, and constipation 53%. Acrocyanosis or vasomotor disorders were present in 16 patients, blood count anomalies in 12, and hypothyroidism in seven.

**Table 1.**
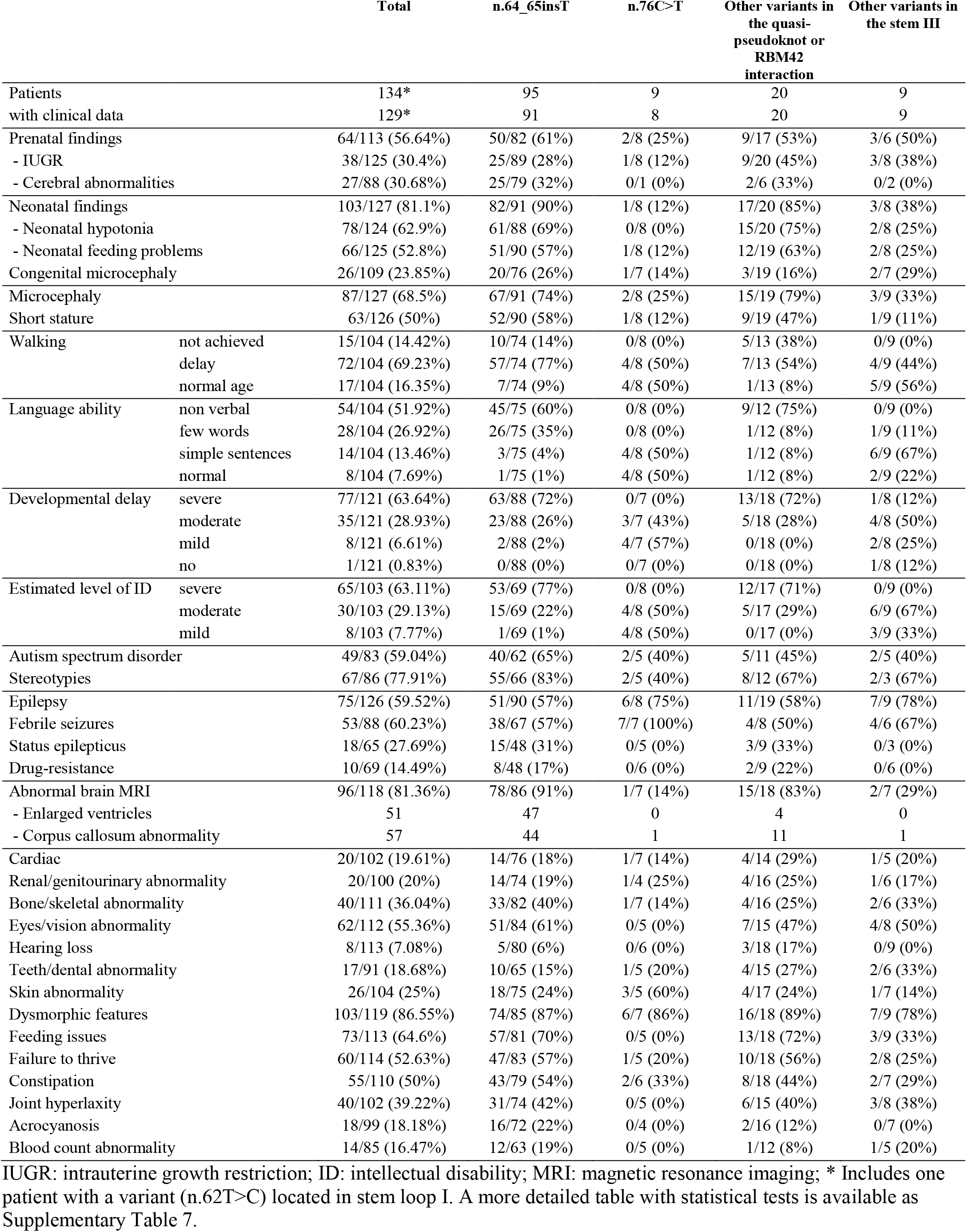
Clinical features of individuals with *RNU4-2* variants according to the location of the variants in the different U4 functional domains.

The phenotype of the patients with variants in U4 T-loop and RBM42 interaction region was indistinguishable from that of patients with the n.64_65insT patients. Patients with the recurrent n.66A>G (*n*=5) and n.67A>G variants (*n*=5) had a similar phenotype, characterized by neonatal hypotonia (5/5 and 3/5), microcephaly (5/5 for both variants), epilepsy in about half of them (3/5 and 2/4), and similar dysmorphic features. All patients had severe developmental delay and severe ID, except for one case with moderate ID. Notably, all patients were non-verbal.

Patients with *RNU4-2* n.76C>T variant (*n*=8) exhibited a distinct clinical profile from patients with the n.64_65insT (Table 1; Supplementary Table 7). They had less neonatal findings (Fisher’s *p*=6.58E-04), specially hypotonia (*p*=0.0208), presented less severe ID (*p*=1.06E-06) and developmental delay (*p*=6.63E-04), were more proficient in their language abilities (*p*=1.06E-06), and rarely showed brain MRI abnormalities (*p*=2.97E-03). All patients could walk, four of them achieving this milestone at a normal age (median walking age: 19m [12-33m]), and all could speak, with simple sentences (*n*=4) or normal language skills (*n*=4). IUGR was observed in only 1/8 patients. Microcephaly was noted in 2/8 patients, and short stature in 1/8. Two out of five patients had autistic features. Six patients had fever-sensitive generalized epilepsy, well-controlled with anti-seizure medication, while two others had a single febrile seizure. None had nystagmus or ataxia, and brain MRI was normal in 5/6 cases, with one showing moderate diffuse cortico-subcortical atrophy. Congenital malformations were rare, and the dysmorphic features were distinct from those seen in patients with the recurrent variant.

Similarly, patients with other variants in the stem III (*n*=9) exhibited a mild/moderate phenotype than patients with the n.64_65insT variant showing less severe intellectual disability (*p*=1.52E-04) and developmental delay (*p*=3.10E-02), and with improved language abilities (*p*=7.79E-06). All patients could walk and speak, with varying degrees of language development (2: normal language, 6: simple sentences and 1: few words). ID was mild in three and moderate in six, with autistic features in 2/5 cases. Fever-sensitive epilepsy was common (7/9), but well controlled with anti-seizure medication. Brain MRI was normal in 5/7 patients.

### *RNU5* variants are associated with NDD with variable malformations

Detailed clinical data were available for 9 out of 15 NDD patients with *RNU5B-1* LP variants (Supplementary Fig. 8; Supplementary Table 8). Six had severe developmental delay, one had moderate developmental delay, and one had normal cognition but attention difficulties. All nine patients showed brain MRI abnormalities, but only one had epilepsy. Three had pectus excavatum, two of whom also had marfanoid habitus. Three had ocular abnormalities such as congenital glaucoma (*n*=1), small papillae with retinal vascular tortuosity (*n*=1) and severe myopia (−15.75/-12.25 dioptries). Other malformations included pulmonary issues (*n*=2), sacrococcygeal abnormalities (*n*=2), tooth agenesis or fusion (*n*=2), and cardiac malformation (*n*=2). Acquired microcephaly was noted in three individuals with n.44A>G, whereas two subjects with n.39C>G had macrocephaly. HPO terms enriched in *RNU5B-1* cases from GEL include seizures, macrocephaly and dystonia (Supplementary Table 9).

The three patients with *RNU5A-1* variants also had NDD with variable congenital malformations. One had postaxial polydactyly, dental agenesis, and talus feet due to oligohydramnios. Another had anal malposition, sacrococcygeal dimple and caudal appendix, thin and incomplete corpus callosum, and septal agenesis; the third had cardiac malformations and marfanoid habitus. The two patients with n.40_41insA had seizures. Head circumference was normal in all.

### Pathogenic variants in *RNU4-2* lead to alternative 5’ splice site usage

Chen et al. reported specific alternative 5’ splice site (5’SS) abnormalities in the blood of individuals with *RNU4-2* variants.^13^ To confirm and extend this observation, we performed RNA sequencing on short-term lymphocyte cultures from 19 *RNU4-2* affected individuals, (10 with n.64_65insT and 9 with other variants) and 21 control individuals with other NDDs. Using rMATS^20^ with an FDR of 0.1 and an absolute deltaPSI difference > 0.05, we detected 35 significant aberrant splicing events affecting 3’ splice sites (3’SS), 126 for mutually exclusive exons, 100 for intronic retention, 121 for exon skipping and 111 for 5’SS (Supplementary Tables 10-14). We then extracted PSI values of significantly altered exons for each splicing category and we performed a principal component analysis using matrices with samples as columns and PSI values as rows. We observed no separation between affected and control individuals for 3’SS and intronic retention, and only a slight separation for mutually exclusive exons and exon skipping (Supplementary Fig. 9). The most striking effect was for 5’SS. We observed a cluster of patient samples with severe phenotypes (c.64_65insT, n.67A>G, n.68A>C and n.70T>C). Patients with a mild phenotype appeared intermediate between severe cases and controls (n.72_73del, n.75C>G, n.76C>T) (Fig. 4A). These results suggest a common signature of 5’SS usage in *RNU4-2* probands, with distinct profiles correlating with disease severity.

**Figure 4.**
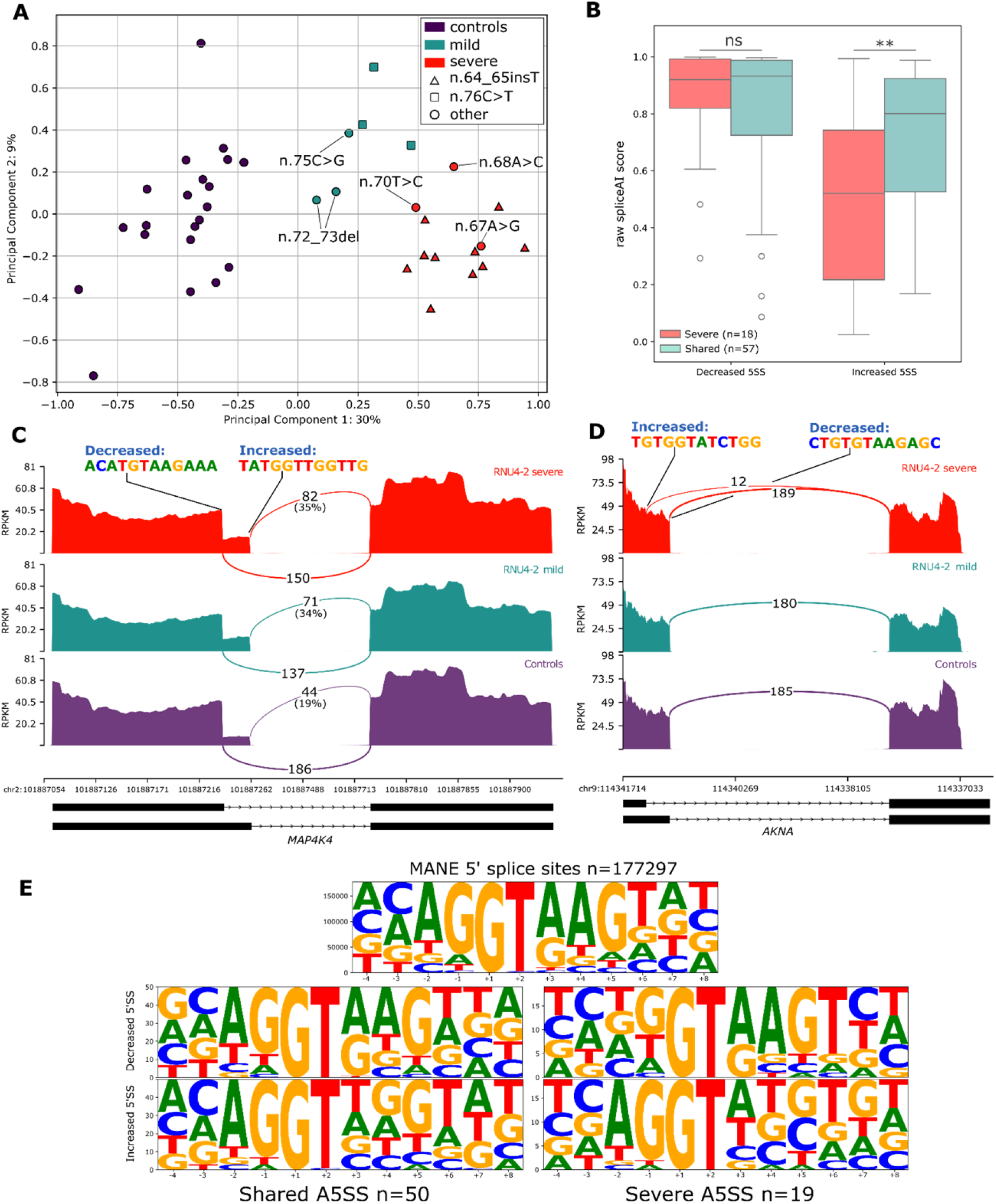
RNA-Seq identifies an alternative 5’SS pattern which differentiates severe from mild *RNU4-2*-related phenotypes. **(A)** Principal component analysis using PSI values of significant 5’SS detected using rMATS (*n*=111). Colors distinguish controls (purple) from mildly affected (green) and severely affected individuals (red). **(B)** Box plot showing raw spliceAI scores of the decreased 5’SS site and the increased 5’SS, for the events shared between mild and severe individuals and for those only detected in severe individuals. SpliceAI scores for severe and shared 5’SS were not statistically different for decreased sites (*p*=0.62) but were significant for increased sites (*p*=0.005) using t-test. Box plot elements are defined as follows: center line: median; box limits: upper and lower quartiles; whiskers: 1.5× interquartile range; points: outliers. **(C)** Sashimi plot showing aggregated coverage and splicing-supporting reads for ‘mild’ *RNU4-2* variants (n.75C>G, n.76C>T, n.72_73del), severe variants (n.64_65insT, n.67A>G, n.68A>C, n.70T>C) and controls, on *MAP4K4* gene. **(D)** Sashimi plot showing on *AKNA* the usage of a 5’SS only for severe variants. **(E)** Consensus nucleotide sequence of decreased and increased 5’SS for 50 shared events (left) and for the 19 severe-only events (right), in comparison to the consensus sequence of all 5’SS from MANE transcripts (top).

We visually characterised 69 5’SS events using *Integrative Genomics Viewer* (IGV), including 50 shared by patients with mild and severe phenotypes and 19 unique to severe phenotypes (Fig. 4B-D; Supplementary Table 15; Supplementary Fig. 10). Decreased 5’SS consistently show high spliceAI scores (shared sites, median=0.93; severe-only sites, median=0.92), indicating that alternative 5’SS usage is not restricted to weak sites (Fig. 4B). In contrast, increased 5’SS events associated with severe phenotypes have significantly lower SpliceAI scores (median=0.52, t-test *p*=0.005) compared to shared sites (median=0.80). Additionally, only five 5’SS events were absent from controls (mean supporting reads in controls <3; 5/69; 7.2%), suggesting that the main effect of U4 variants is a shift in existing alternative isoforms rather than the use of new cryptic splice sites. The consensus of 5’SS usage revealed a consistent decrease in A/A/G nucleotides replaced by C/T at positions +3/+4/+5 (Fig. 4E). This effect is associated with an increase in the dependence of A/G in position -2/-1 that is particularly visible for decreased 5’SS events restricted to severe phenotypes (2/19 ‘AG’ for severe variants vs. 26/50 ‘AG’ for shared variants; 2-tailed Fisher’s test, *p*=0.0008). This suggests that 5’SS used only in severe patients are more reliant on the end of the exon and less on the sequence of the intron. Altogether, we identified a specific splicing pattern associated with *RNU4-2* variants that can distinguish affected samples based on disease severity and variant location.

### Identification of a specific *RNU4-2* episignature

Finally, we investigated whether *RNU4-2* variants are associated with specific DNA methylation profiles. We analysed methylation profiles of 23 patients with P/LP variants in *RNU4-2* and 35 controls (healthy, age-matched individuals). Following adjustment on age, sex and blood cell composition, epigenome-wide analysis led to the identification of 90 probes with uncorrected p-value < 10^−7^ and |Δβ| > 5%. PCA and heatmap representations of adjusted methylation levels showed a good separation between moderate-to-severe *RNU4-2* phenotypes and controls while mild NDD phenotypes tend to cluster with the controls (Fig. 5). The first PC axis, separating moderate to severe patients from controls captures 54% of the residual variance of methylation levels, after adjustment for age, sex and cell composition (Supplementary Fig. 11), which is of the order of magnitude previously observed for *ATRX, KMT2D* or *KMT2A* episignatures.^21^ After five-fold cross-validation, the signature obtains a sensitivity of 0.68 (95% CI [0.47 – 0.85]), and a specificity of 0.97 (95% CI [0.85, 0.999]) (Supplementary Fig. 12).

**Figure 5.**
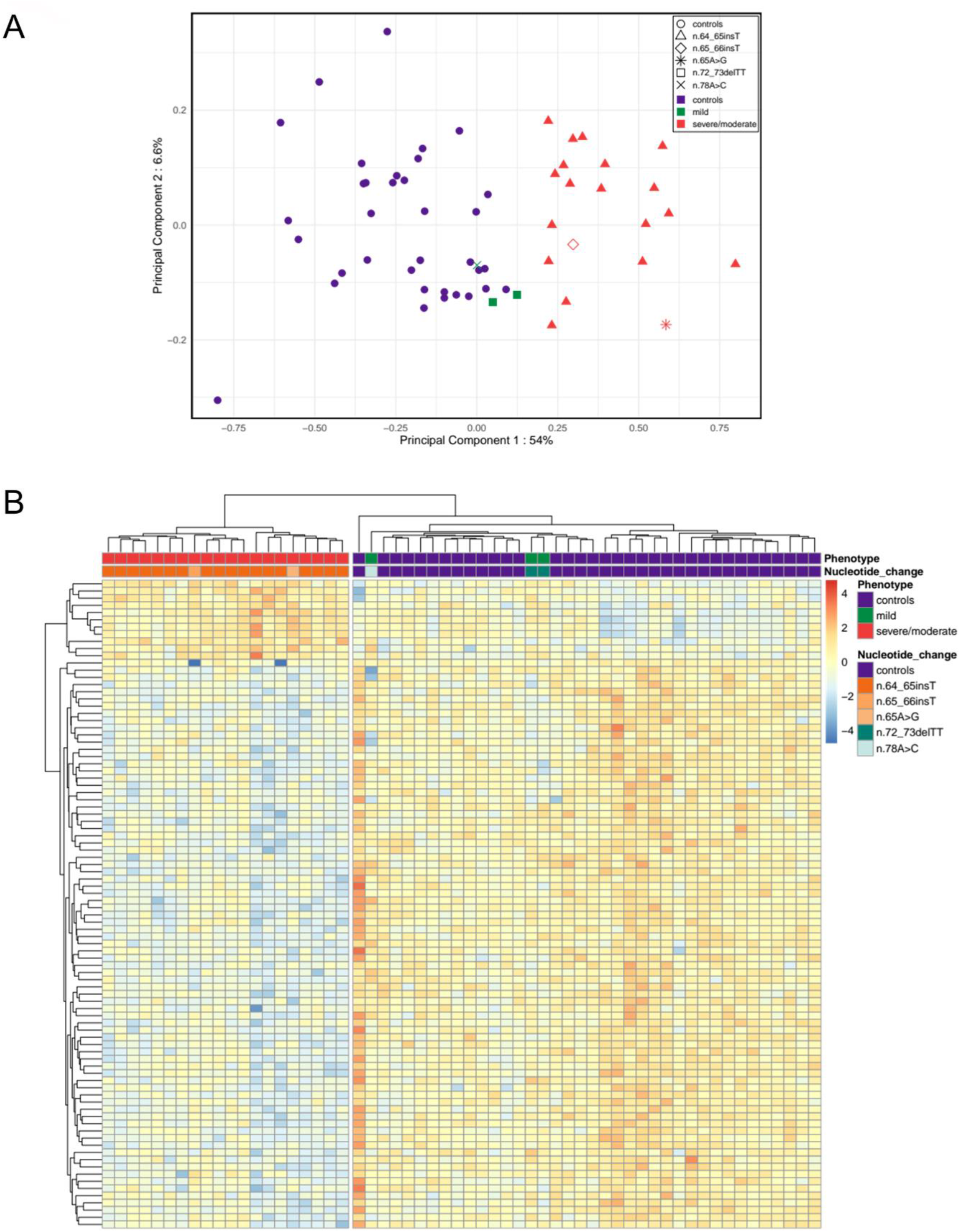
*RNU4-2* episignature differentiates moderate-to-severe NDD phenotypes from controls. **(A)** Principal component analysis of residual methylation levels after adjustment for age, sex and estimated blood cell counts. Different variations are represented by different shapes. Phenotypes are distinguished by color, blue for controls, green for mild phenotypes, red for moderate to severe phenotypes. Percentage of explained variance is added to each axis. Heatmap of residual methylation levels displays hierarchical clustering of controls and *RNU4-2* patients. Blue indicates hypo-methylated positions while red indicates hyper-methylated positions with respect to expected methylation levels at equivalent age, sex and blood cell composition.

## Discussion

Despite extensive genetic testing, 40-60% of patients with NDDs of suspected genetic origin remain unsolved. This diagnostic gap is due to several challenges including difficulties in interpreting variants in non-coding genomic regions. The recent discovery of *RNU4-2* variants as a major cause of NDDs, overlooked until 2024, highlights the significant role that non-coding genes likely play in undiagnosed cases. Here, we analysed genes encoding functional snRNAs in a large French cohort of patients who underwent genome sequencing as part of routine diagnosis and identified 74 patients with pathogenic or likely pathogenic variants in *RNU4-2* (0.49% of subjects with NDD) and seven (0.046%) with variants in the 5’ loop I of *RNU5A-1* or *RNU5B-1*. We also collected data of 73 additional patients from distinct cohorts. Altogether, we observed that pathogenic variants cluster in evolutionarily conserved, key functional regions of U4 and U5 involved in splicing. Specifically, *RNU4-2* variants cluster in two main U4 regions, the T-loop/quasi-pseudoknot and stem III, while variants in *RNU5B-1* all are in the conserved loop I, the part of U5 that pairs with the end of the exon adjacent to the 5’SS.^5^ The three *de novo* variants in *RNU5A-1* are also located in this constrained region. A few additional *de novo* variants in other domains or snRNA genes were also identified but their clinical relevance remains unclear.

This study provides a comprehensive overview of *RNU4-2-*related phenotypes based on a large series of patients, and shows distinct clinical outcomes depending on variant location. Variants in the T-loop, including n.64_65insT, are associated with more severe phenotypes, while variants in the stem III lead to milder forms. The n.76C>T variant, previously suggested to cause a milder phenotype from single case observations^13,14^, and c.72_73del, support the idea of a continuum of *RNU4-2*-related phenotypes, with inherited variants also possibly contributing to the genetic aetiology of NDDs.

We observed prenatal manifestations in 60% of individuals with *RNU4-2* pathogenic variants, mainly isolated cerebral abnormalities (corpus callosum anomalies and enlarged ventricles) and/or IUGR. Considering the high incidence of *RNU4-2* variants, these findings have noteworthy implications for prenatal diagnosis, emphasising the need for genome sequencing, or adding *RNU4-2* analysis, in prenatal genetic testing.

A striking observation in line with previous findings^13^ is the predominant maternal origin of *RNU4-2* variants, possibly explained by negative selection of variants severely affecting splicing in the male germline. However, paternal transmission of less severe variants is possible, as evidenced by three cases. The mechanism underlying the high recurrence of *RNU4-2* and its potential link to maternal origin remain unclear. Interestingly, recurrent insertions in *RNU4-2* n.64_65insT and *RNU5A-1* n.40_41insA occur at 2’-O-methylation sites,^11,12^ though any connection to maternal inheritance or recurrence is yet to be established.

We provide definitive evidence that *RNU4-2* pathogenic variants lead to specific alternative 5’SS anomalies in blood cells of affected patients, with detected events correlating with phenotype severity. Variants in the T-loop and stem III indeed show distinct, partially overlapping transcriptional signatures, which could be used to interpret variants of uncertain significance. Furthermore, the more severe NDD phenotypes associated with *RNU4-2* variants in the T loop also display a specific methylation profile (episignature). Given the widespread use of exome sequencing in routine diagnostics, these transcriptional and epigenetic signatures could help diagnose additional ReNU syndrome cases worldwide.

From a molecular standpoint, U4 variants may disrupt spliceosome function at various stages: U4 snRNP biogenesis, U4/U6 di-snRNP, U4/U6.U5 tri-snRNP assembly, or spliceosome activation. However, considering their location and associated transcriptional alterations, they more likely disrupt the organization of the U4/U6 duplex at the tri-snRNP stage and affect the introduction of the 5’SS into the spliceosome’s active site. The 5’SS is initially paired with U1 in the pre-spliceosome and then transferred to the U6 ACAGAGA box and U5 stem loop 1 located in U4/U6.U5 tri-snRNP. These interactions maintain the 5’SS in the active site during catalysis, marking the start of each intron^1,22^. When the 5’SS is transferred, it pairs with the U6 ACAGAGA box to ensure its correct identification ^23^. Proper pairing triggers molecular events that lead to the formation of the active site by Brr2. Before this transfer, the U6 ACAGAGA box is held as a flexible loop between the stem III and quasipseudoknot. Snu66, SNRP27K, and RBM42 proteins further stabilize this organization, ensuring the ACAGAGA box can properly recognize the 5’SS.^4,24-26^

Pathogenic *RNU4-2* variants fall under two main categories. Variants in -or close to-the quasi-pseudoknot (n.62-67) possibly weaken if not disrupt its structure and compromise its ability to maintain the ACAGAGA box at the right position for 5’SS recognition. A similar effect could apply to variants (n.68-70) located in nearby single stranded U4 region interacting with RBM42. The second variant category (n.73-79) likely affects U4/U6 stem III’s stability by altering Watson Crick base pairs formed by U4’s A78, C76 and C75 with U6’s G34, G33 and U31. The role of stem III in 5’SS recognition remains unclear. Besides orienting the ACAGAGA box, stem III may enhance fidelity by imposing an energy barrier to the extension of a U6/5’SS helix stemming from the initial pairing of the 5’SS with the ACAGAGA sequence^4,5^. Stem III disruption is required prior to Brr2 loading onto U4 for active site formation. Hence, we hypothesize that a weakened stem III may allow sub-optimal 5’SS to more easily extend the U6/5’SS helix, promoting spliceosome activation. The increased 5’SS in the severe cases suggests that the spliceosome loses specificity for the intronic 5’SS sequence motif, which seems to be compensated by more contribution from the exonic 5’SS sequence motif by U5.

U5 loop I plays an important role during 5’SS transfer by interacting with the exonic sequences adjacent to 5’SS. These interactions help alignment of the 5’exon with respect to the branch site and the 3’exon during both steps splicing catalysis^27,28^. Mutations in yeast U5 loop I result in the activation of the aberrant 5’SS, making it an important determinant of 5’SS specificity^29^. *RNU5* variants may therefore modify the specificity of U5 loop I towards pre-mRNA, resulting in similar effects as *RNU4-2* variants. Future studies should analyse whether there is a similar or distinct splicing signature associated with variants in *RNU5A-1* and *RNU5B-1* to that present in *RNU4-2*. Together, these RNA structures formed by U4 and U5 build a conformation that prevents weak 5’SS-triggered activation of the spliceosome, promoting the usage of optimal 5’SS. Destabilization of these structures would make the spliceosome more prone to error, leading to a decrease in splicing fidelity.

In conclusion, this work emphasises the critical role of *de novo* variants in snRNAs, particularly *RNU4-2*, in unsolved NDD. Moreover, we identify *RNU5B-1* as a novel NDD gene, and provide valuable insights into fundamental aspects of spliceosome function.

## Methods

### Inclusion & Ethics statement

This study complies with the ethical standards of each of the participating countries. An informed consent was obtained for all patients included in this study from their parent or legal guardian. A specific consent form was obtained from the families who consented to publication of photographs. Patient/participant/samples were pseudonymized for the genetic study at each participating center. We collected information on the sex (but not gender) of the patients from the patients’ clinical file. The study has received the approval of the ethics committee of University Hospital Essen (24-12010-BO). For methylation analysis, DNA from all individuals (patients and controls) had been collected previously in the context of genetic analysis in a medical setting, following signature of a written, informed consent that includes a query on the use of leftovers in a research setting. Healthy controls consisted in individuals without NDD who underwent pre-symptomatic testing for other conditions and were found to be non-carriers or unaffected relatives of patients with a genetic disease, among non-carriers of pathogenic variants. Samples used for the methylation study were stored within the genetics biological collection of the CRBi, Rouen, France, declared as DC 2008-711 (access authorization n° MCRBi/2024/02). The analysis of methylation profiles based on previously stored DNA in these conditions was approved by the CERDE ethics committee (notification n° E2023-13) from the Rouen University Hospital. Researchers and clinicians from participating centers contributing either data or intellectual input were involved at all stages of the study from design, implementation, drafting, and revising the manuscript, and are coauthors of the article.

### List of snRNA genes

A list of 50 official gene symbols encoding functional snRNAs (Supplementary Table 3) was established from the HUGO gene nomenclature committee (https://www.genenames.org/). Information was retrieved from HGNC in December 2023 by applying the advanced filtering “gd_locus_type = ‘RNA, small nuclear’“ and restricting to genes with approved symbols.

### Patient cohorts

We initially identified the n.64_65insT variant in a single patient with developmental epileptic encephalopathy. This variant was prioritized because it was the strict *de novo* variant with the highest CADD score and was submitted to GeneMatcher^30^. Following the publication of the preprint by Chen et al. on April 8, 2024^31^, we investigated variants in *RNU4-2* and 49 other snRNA genes in several diagnostic and research cohorts. Our inclusion criteria were: 1) a *de novo* variant in any of the 50 snRNA genes with less than 10 heterozygotes in gnomADv4.1.0, or 2) a heterozygous variant in *RNU4-2* located within the critical 18-nucleotide region as defined by Chen et al. 2024.^13^ We then narrowed our search to *RNU5A-1* and *RNU5B-1*, and investigated 3) *de novo* variants with less than 10 heterozygotes in gnomADv4.1.0 and/or 4) heterozygous variants in *RNU5B-1* located in the 5’ loop I.

The main cohort is composed of 23,649 patients with rare disorders, including 15,073 patients with NDD and their parents, when available, who underwent genome sequencing as part of the diagnostic process in France (Plan France Médecine Génomique 2025, PFMG2025) on one of the two national clinical sequencing laboratories, SeqOIA (https://laboratoire-seqoia.fr/) and Auragen (https://www.auragen.fr/) between 2019 and 2024. All *de novo* variants were visualized on IGV. The analysis of *RNU4-2* variants in this main cohort identified 78 patients. Furthermore, we collected data of patients with *de novo* and/or pathogenic *RNU4-2* variants identified in either diagnostic or research contexts through national networks, established collaborations, or GeneMatcher^30^. These additional cohorts included 40 patients from France, 17 patients from Germany, one patient from the Netherlands, one from Spain, and one from the US. Twenty-two patients had genome sequencing in diagnostic (*n*=5) or research (*n*=15) contexts, whereas in 38 patients, the variant was identified or confirmed by a targeted method: Sanger sequencing (*n*=31) or next generation sequencing of amplicons (*n*=5). Among the patients who were diagnosed by Sanger sequencing, two patients previously had inconclusive exome analysis and were included in SOLVE-RD. Reads supporting the presence of n.64_65insT were identified in the exome data. None of the patients included in this study had been previously published, and we also checked that there were no duplicates for individuals with the same variant based on the individual’s year of birth and initials.

The analysis of *de novo* variants in the other 49 snRNA genes in the PFMG cohort identified six patients with *de novo* variants *RNU5B-1*, three patients with *de novo* variants in *RNU5A-1*, and six patients with *de novo* variants in a single gene: *RNU4-1, RNU5E-1, RNU5F-1, RNU6atac, RNVU1-22*, or *RNVU1-27* (Supplementary Table 4). A targeted search for variants in *RNU5B-1* and *RNU5A-1* in the Genomics England dataset (including both the 100,000 Genomes cohort (v18) and National Health Service Genomic Medicine Service (NHS-GMS v3) cohort) identified six additional individuals with rare (< 10 occurrences in gnomAD) *de novo* variants, five out of 8,841 undiagnosed NDD proband and one out of 21,816 non-NDD probands. In addition, three probands analysed in duo had a rare variant in the 5’ loop I absent from the single parent analysed. In addition, five *de novo* variants in *RNU5B-1* were collected from the Broad Centre for Mendelian Genomics, Undiagnosed Disease Networks (UDN), the BCH Epilepsy Genetics Program, and Care4Rare Canada.

### Variant classification

We classified variant according to ACMG/AMP criteria^18^ using recommendations from Ellingford et al.^19^ The PS2 (or PM6 for patients who underwent targeted sequencing) criteria was applied for all cases with *de novo* inheritance. We applied PS3 when RNAseq and/or methylation analyses supported pathogenicity or PM1 for variants located in mutational hotspots: chr12(hg38):120,291,825-120,291,842 for *RNU4-2* and chr15(hg38):65,304,713-64,304,720 for *RNU5B-1*. PM2 supporting was applied for variants absent from gnomAD v4.1.0, and PS4 supporting was applied for recurrent variants (at least 3 patients in this study or in Chen et al.^13^). This led to classify *de novo* variants located in the critical region as LP. Recurrent variants with > 3 occurrences and absent from databases were classified as P. Only variants for which inheritance was unknown (even located in critical region) or variants occurring outside of critical regions (even if *de novo*) were classified as variants of uncertain significance (VUS).

### Clinical data analyses

Clinical data were retrospectively collected from the referring physician using an anonymized Excel sheet. Categorical data for 44 selected clinical features from 129 patients with pathogenic and likely pathogenic *RNU4-2* variants were converted to a 0-1 scale, with 0 representing a more favourable phenotype presentation and 1 a more severe phenotype. Hierarchical clustering was performed using pheatmap R package, performing Z-score scaling for each row (across different patients), and ward.D2 clustering method keeping missing values. PCA was generated after replacing missing data by 0 and performing variable scaling. Plots concerning to head of circumference (HC) measurements at birth were generated with the “Plotter: Preterm growth charts, 22-50 weeks” from the Canadian Pediatric Endocrine Group (https://cpeg-gcep.shinyapps.io/prem2013/).^32^ For additional HC measurements, reference chart data points were obtained from Rollins et al. 2010.^33^ Male patients older than 21 years were plotted at age 21 and female patients older than 20 years were plotted at age 20, corresponding to the maxima for each sex. Fisher’s tests (two-sided; 2×2, 2×3 or 2×4 contingency tables) adjusted for multiple comparisons using Bonferroni correction were used to compare clinical features in different U4 domains (n.64_65insT vs. n.76C>T and n.64_65insT vs. the other variants groups) for the 39 clinical features.

### Conservation and *in silico* predictions

The highest homolog to the human *RNU4-2* and *RNU5B-1* were obtained for *Ciona intestinalis, Ciona savignyi, Drosophila melanogaster, Caenorhabditis elegans, Danio rerio* and *Mus musculus* by using BLAT on each of these genomes in Ensembl Release 112.^34^ RNA sequences from *RNU4-2* and *RNU5B-1* were aligned to i) their respective sequence homologs and 2) the sequence(s) of other U4- and U5-encoding genes expressed in the brain using Geneious Prime® 2019.2.3 (Geneious Alignment). The threshold for consensus was set to 100% identical, highlighting positions with 100% agreement between all sequences.

CADD PHRED scores and conservation in vertebrate (verphyloP) were calculated for pathogenic and likely pathogenic patient variants and gnomADv4.1.0 variants with CADD v1.7^35^. For each variant, in silico-mutated U4 RNA sequences were generated with seqkit mutate^36^. Bifold^37^ was used to generate the multiple U4:U6 interactions and calculate the minimum free energy. Comparisons were performed by applying Mann-Whitney U test, two-sided.

### Expression of snRNAs in brain tissues

We used small-RNA data for different human embryonic brain regions to inspect the expression level of selected snRNAs. These data were generated by the ENCODE Consortium^38^: diencephalon (GEO:GSE78292), temporal lobe (GEO:GSE78303), occipital lobe (GEO:GSE78298), frontal cortex (GEO:GSE78293), parietal lobe (GEO:GSE78299), cerebellum (GEO:GSE78291). Tracks show unique read signals for plus and minus strand from the default anisogenic replicate. Expression of these genes in the brain using BrainVar was previously investigated.^13^

### RNA-sequencing

Peripheral blood mononuclear cells (PBMCs) were isolated from 2 to 4 ml of EDTA-anticoagulated blood within 48 hours of collection using UNISEP+ tubes (EUROBIO). Cells were cultured in 6-well plates (5.0×10^5^ to 2.0×10^6^ cells per well) in lymphocyte-stimulating medium (Chromosome Medium P, EUROCLONE) for 48 to 72 hours at 37°C (5% CO2). After incubation, one well per sample was treated with 1mg/ml puromycin solution for 4-5 hours to inhibit Nonsense-Mediated Decay (NMD). RNA was extracted using the NucleoSpin RNA Plus extraction kit (MACHEREY-NAGEL) according to the manufacturer’s instructions.

Stranded RNA-Seq libraries were prepared from 100ng of total RNA on the Magnis NGS Prep System (Agilent) using SureSelect XT-HS2 kit (Human All Exon V8 capture probes) with 12 and 10 PCR-cycles for pre-capture and post-capture amplifications, respectively. RNA-Seq were sequenced on an Illumina’s NextSeq 550 (16 samples on HighOutput 2×75bp) to obtain 25 to 30 million paired-end reads per sample.

Fastq files were aligned on GRCh38 reference genome with STAR (v.2.7.11a), in two-pass mode, using Ensemble transcripts v.106. Quality control was performed with fastqc (v.0.11.3) and fastp (v.0.23.4). Controls were matched on following criteria: library preparation kit, sequencing flowcell and culture time. CIBERSORTx was used to estimate relative abundance of blood cells using LM22 signature matrix file^39^. One *RNU4-2* sample was removed because of a low proportion of activated T CD4+ cells (1/38). Abnormal splicing events were called using 19 *RNU4-2* samples and 21 controls by using rMATS (v.4.3.0)^20^ with following parameters: -t paired –anchorLength 1 –libType fr-firststrand –task both –novelSS – variable-read-length –allow-clipping. Python scripts were used to filter rmats output files with following filters: mean coverage > 7, FDR < 0.1, deltaPSI > 0.05. Principal component analysis was performed using sklearn python library using PSI values from significant A5SS, keeping only the most significant A5SS if several were called in the same exon. Raw spliceAI scores were obtained from MobiDetails^40,41^.Sashimi plots were made using rmats2sahimi and boxplots with seaborn. Consensus nucleotide sequences were generated using Logomaker.

### Epigenome-wide analysis and DNA methylation signature

Genomic DNA was extracted from whole blood and bisulfite converted. DNA methylation profile was then derived using Illumina’s Infinium EPIC array v2.0, in accordance with the manufacturer’s protocol. Patients and negative controls were balanced across sixteen arrays and each array rows in order to reduce technical biases. DNA methylation arrays were generated at the ASGARD-Rouen genomic platform (University of Rouen and Rouen University Hospital, Rouen France) on an Illumina NextSeq550 scanner. Raw IDAT data were processed and normalised using the default Meffil R package protocol along with all other samples included in the sixteen arrays in order to better estimate the variability of methylation signals within and across arrays^42^. One *RNU4-2* sample failed default quality controls and was excluded from further steps. Remaining samples were functionally normalised together as advocated in the Meffil documentation, with random effect adjustment on array and sentrix row as well as fixed effect adjustment on the first two PCs, before computing β-values.

Several predictions were obtained from methylation values to apply additional QC and normalization steps. Sex predictions were extracted from the standard Meffil normalized object. No inconsistencies between reported and predicted sex were noted (Supplementary Fig. 11A). Blood cell counts were estimated with the meffil.cell.count.estimates function. PCA of predicted blood cell counts showed a good overlap of positive and negative controls in terms of overall blood cell composition (Supplementary Fig. 11B). DNA methylation age was predicted with the DNAmAge function from the methyclock R package^43^. The Horvath and skinHorvath clocks both displayed very strong correlation with actual age at blood sample on our dataset (Pearson correlation *r*=0.97 (Supplementary Fig. 11C-D).

The set of differentially methylated probes was identified with the meffil.ewas function on the subset of controls and pathogenic or likely pathogenic *RNU4-2* variant carriers. To correct for well-known confounders, the analysis was adjusted on age at blood sample, sex and predicted blood cell composition. Manhattan and volcano plots are given in Supplementary Fig. 12A-B. After filtering on p-value and average methylation difference between positive and negative controls, residuals of a linear regression model fitted to adjust probe β-values on age, sex and predicted cell composition, were visualized through principal component analysis and heatmap representations. Phenotype classification into mild/moderate and severe subtypes was derived independently from the episignature discovery and a posteriori added to these graphical representations.

Finally, the robustness of the signature was challenged through 5-fold cross-validation. The dataset was split into five random and equal-sized blocks. Each block was used in turn as validation set, while the remaining four blocks were used as training set to run a new differential analysis based on controls and moderate to severe phenotypes. An SVM model was trained on each cross-validation training set, and applied to the test set to derive unbiased sensitivity and specificity estimations overall, and by phenotype class, along with 95% binomial confidence intervals.

### Variant impact

Structural analysis of variants and corresponding figures were performed using the PyMol v3.0.0 visualisation software^44^ on published coordinates of the human tri-snRNP structure: PDB 6QW6^4^ and PDB 8Q7N^45^.

## Data availability

Individual genome, RNAseq and methylation data could not be made publicly available due to ethical considerations. Controlled access to human sequences is necessary to protect the privacy of participants and to ensure that the use of the data conforms to ethical and legal standards, particularly in relation to data protection laws in France, Germany, and Europe. For inquiries regarding data access, please contact the corresponding authors by email. Other data supporting the findings described in this manuscript are available in the article and its Supplementary Information files. We also used data from Ensembl Release 112 and data from ENCODE Consortium: bigwig files with the plus/minus strand signals of unique reads from the default anisogenic replicate from the following tissues: diencephalon (https://www.encodeproject.org/experiments/ENCSR000AFR/), parietal lobe (https://www.encodeproject.org/experiments/ENCSR000AFY/), occipital lobe, (https://www.encodeproject.org/experiments/ENCSR000AFX/), frontal cortex (https://www.encodeproject.org/experiments/ENCSR000AFS/), temporal lobe (https://www.encodeproject.org/experiments/ENCSR000AGD/), cerebellum (https://www.encodeproject.org/experiments/ENCSR000AFQ/).

## Code availability statement

The analyses were conducted using existing software and packages, including: STAR aligner v.2.7.11a, CIBERSORTx, rMATS v.4.3.0, rmats2sahimi, seaborn, Logomaker, Meffil R package, methyclock R package, ggplot2 v3.3.6, pheatmap v1.0.12, stats v4.2.0, factoextra v1.0.7, and PyMol Version 3.0.0. Free energy for RNA secondary structure was calculated using bifold from RNAstructure v6.5 (https://rna.urmc.rochester.edu/RNAstructure.html). Preterm head circumference graphs were plotted using https://cpeg-gcep.shinyapps.io/prem2013/. Custom scripts used for *RNU4-2* RNAseq analysis are available on GitHub using the following link: https://github.com/benjamin-cogne/RNU4-2_transcriptomics (https://doi.org/10.5281/zenodo.13868502).

## Author contribution

Be.Co., Am.Sa., El.Le., and Fr.Le., developed bioinformatics pipelines and performed data analyses. Y.C. and N.W. performed data analyses in the GEL/NHS cohorts. S.L.S. and A.O’D-L contributed data form other cohorts. Cl.Ch. performed structural analysis. Ca.Na., Ju.Th., and Ch.De. conceived the study, performed data analyses, and supervised the project. All other authors contributed molecular and/or clinical data. Ca.Na., Be.Co., Ca.Ch., Cl.Ch. and Ch.De. drafted the article. All authors reviewed and approved the final manuscript.

## Acknowledgments

We thank the patients and their families for their participation in this study. This research was made possible through access to data in the National Genomic Research Library, which is managed by Genomics England Limited (a wholly owned company of the Department of Health and Social Care). We thank P. O’Donovan, Mitra Sato and Zainab Mustafa from Genomics England for their help with Airlock requests. The National Genomic Research Library holds data provided by patients and collected by the NHS as part of their care and data collected as part of their participation in research. The National Genomic Research Library is funded by the National Institute for Health Research and NHS England. The Wellcome Trust, Cancer Research UK and the Medical Research Council have also funded research infrastructure. We thank the Care4Rare Canada Consortium for support of a subset of this work. The Chair in Genomic Medicine awarded to JC is generously supported by The Royal Children’s Hospital Foundation. We also thank Miranda Di Biase, Ashley Wilson, Atteeq Rehman, Amanda Thomas-Wilson, Shruti Phadke, Avinash Vabhyankar from the New York Genome Center for their help with genome analysis and for providing anonymized patient data. A.K. and F.J.K. from Essen, Germany, T.S.B from the Netherlands, K.Õ. from Estonia, A.-S.D.-P., L.F., G.L., J. V-G., M.W. and S.O. from France, and N.R. from Belgium are members of the European Reference Network (ERN) ITHACA whose EU coordinator in A.V. Support for title page creation and format was provided by AuthorArranger, a tool developed at the National Cancer Institute.

