## Supplementary Information for "Dominant variants in major spliceosome U4 and U5 small nuclear RNA genes cause neurodevelopmental disorders through splicing disruption"

### Supplementary Tables

**Supplementary Table 1:** *De novo* variants or LP/P variants in *RNU4-2* identified in this study

**Supplementary Table 2:** CADD, phyloP and bifold

**Supplementary Table 3:** 49 additional small nuclear RNAs with approved HGNC status

**Supplementary Table 4:** *De novo* PFMG variants in other small nuclear RNAs

**Supplementary Table 5:** *De novo* variants and variants absent from single parents in *RNU5A-1* and *RNU5B-1* in GEL and GMS cohorts

**Supplementary Table 6:** *RNU5B-1* variants in the critical region

**Supplementary Table 7:** Clinical features of individuals with *RNU4-2* LP/P variants

**Supplementary Table 8:** Clinical features of individuals with *RNU5A-1* and *RNU5B-1* variants

**Supplementary Table 9:** HPO terms enrichment for *RNU5B-1* cases from GEL

**Supplementary Table 10:** Significant alternative 5’ splice sites (5’SS) events called by rMATS

**Supplementary Table 11:** Significant alternative 3’ splice sites (3’SS) events called by rMATS

**Supplementary Table 12:** Significant mutually exclusive exons called by rMATS

**Supplementary Table 13:** Significant intron retention events called by rMATS

**Supplementary Table 14:** Significant skipped exons called by rMATS

**Supplementary Table 15:** Characterization of significative alternative 5’SS events with spliceAI scores and nucleotide sequence

### Supplementary Figures

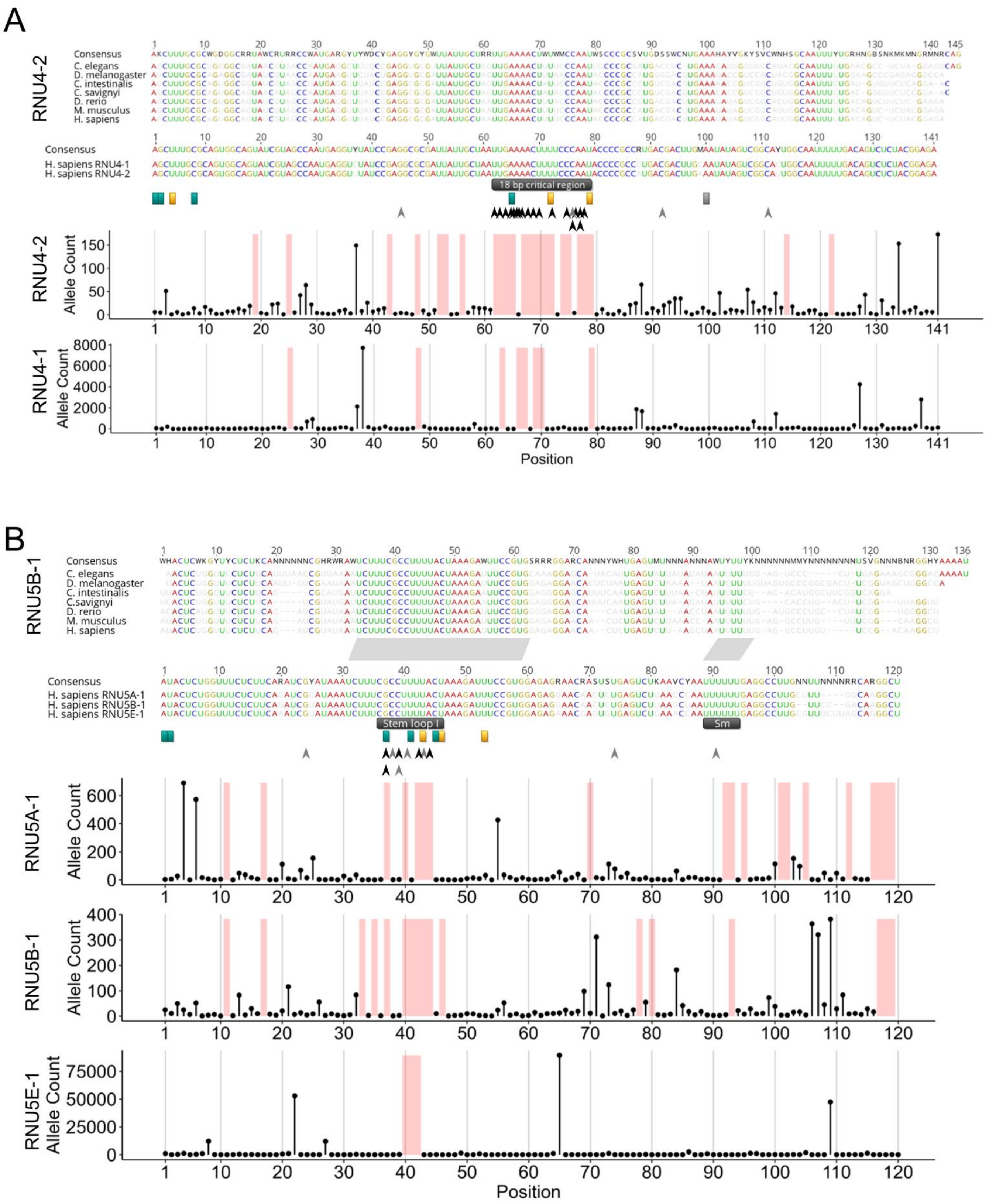


**Supplementary Figure 1: Conservation and constraints of genes encoding U4 and U5 expressed in the brain. (A)** Correspondence between the alignment of *RNU4-2* sequences from animals (top), the alignment of human *RNU4-2* and *RNU4-1* (middle) and the allele counts from *RNU4-2* and *RNU4-1* variants in gnomAD v4.1.0 (bottom). The 18 bp critical region from Chen et al.[13] is highlighted. **(B)** Correspondence between the alignment of *RNU5B-1* sequences from animals (top), the alignment of human *RNU5A-1*, *RNU5B-1* and *RNU5E-1* (middle) and their allele counts in gnomAD v4.1.0 (bottom). The 5’ loop I and Sm regions are highlighted. (**A** and **B**) The threshold for consensus is 100% identical. Nucleotides in red, blue, yellow and green are shown only for positions with 100% agreement between all sequences. Other nucleotides in black (consensus, using also IUPAC codes) or grey (sequences). Arrows indicate variants from this study (pathogenic and likely pathogenic in black, variants of uncertain significance (VUS) in grey). Pseudouridine (yellow), 2‘-O-methyl residues (teal); N6-methyladenosine (grey). Regions shaded in light red are devoid of variants in gnomAD v4.1.0.

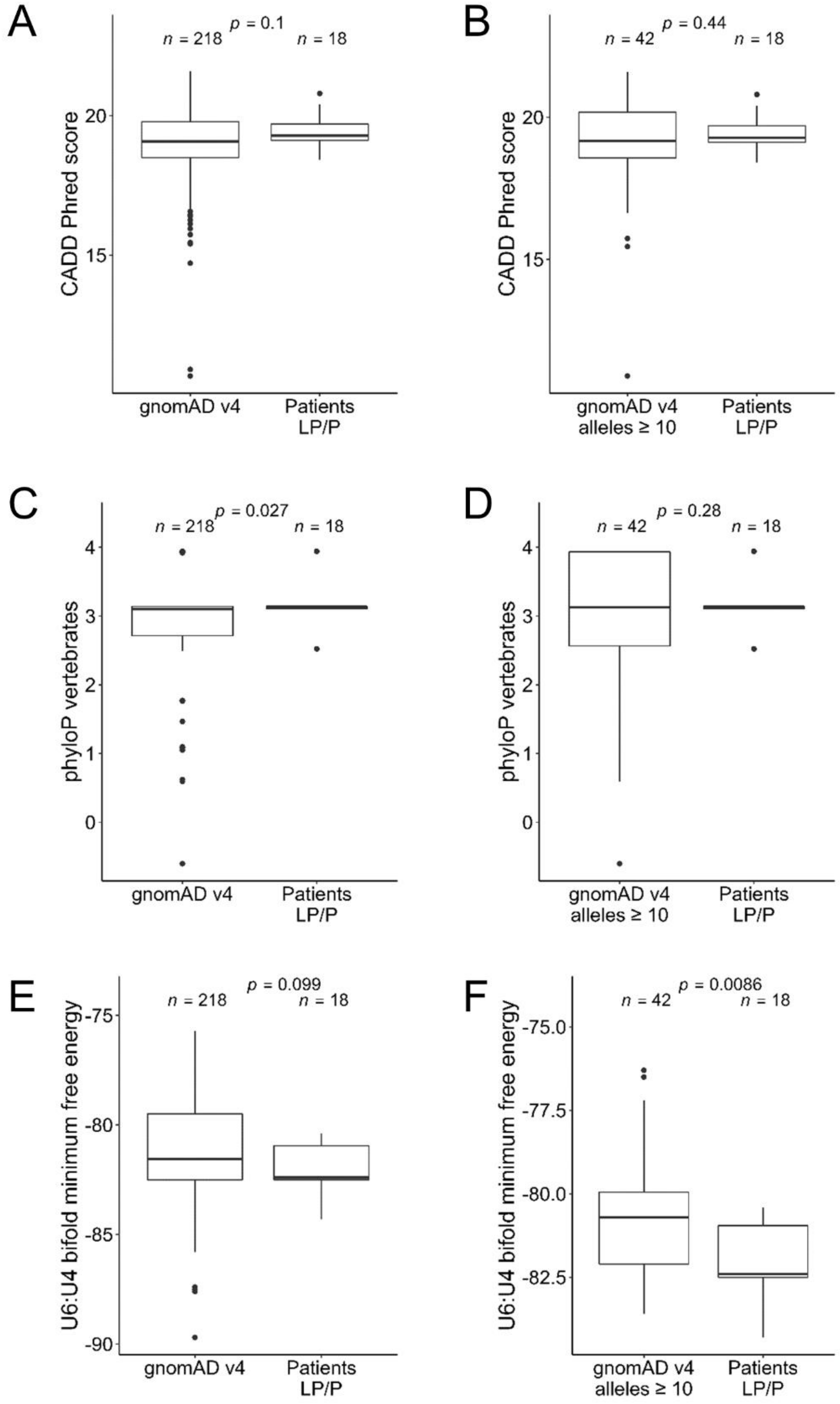


**Supplementary Figure 2: Comparison of features between LP/P variants and variants found in gnomAD.** Distributions of CADD scores (A-B), conservation in vertebrate (phyloP; C-D) and U4:U6 bifold minimum free energy[37] (E-F) for pathogenic and likely pathogenic variants versus gnomAD v4.1.0 variants (A, C and E) or versus recurrent gnomAD v4.1.0 variants (with at least 10 allele counts; B, D and F). Box plot elements are defined as follows: center line: median; box limits: upper and lower quartiles; whiskers: 1.5× interquartile range; points: outliers. Comparisons were performed by applying Mann-Whitney U test, two-sided.

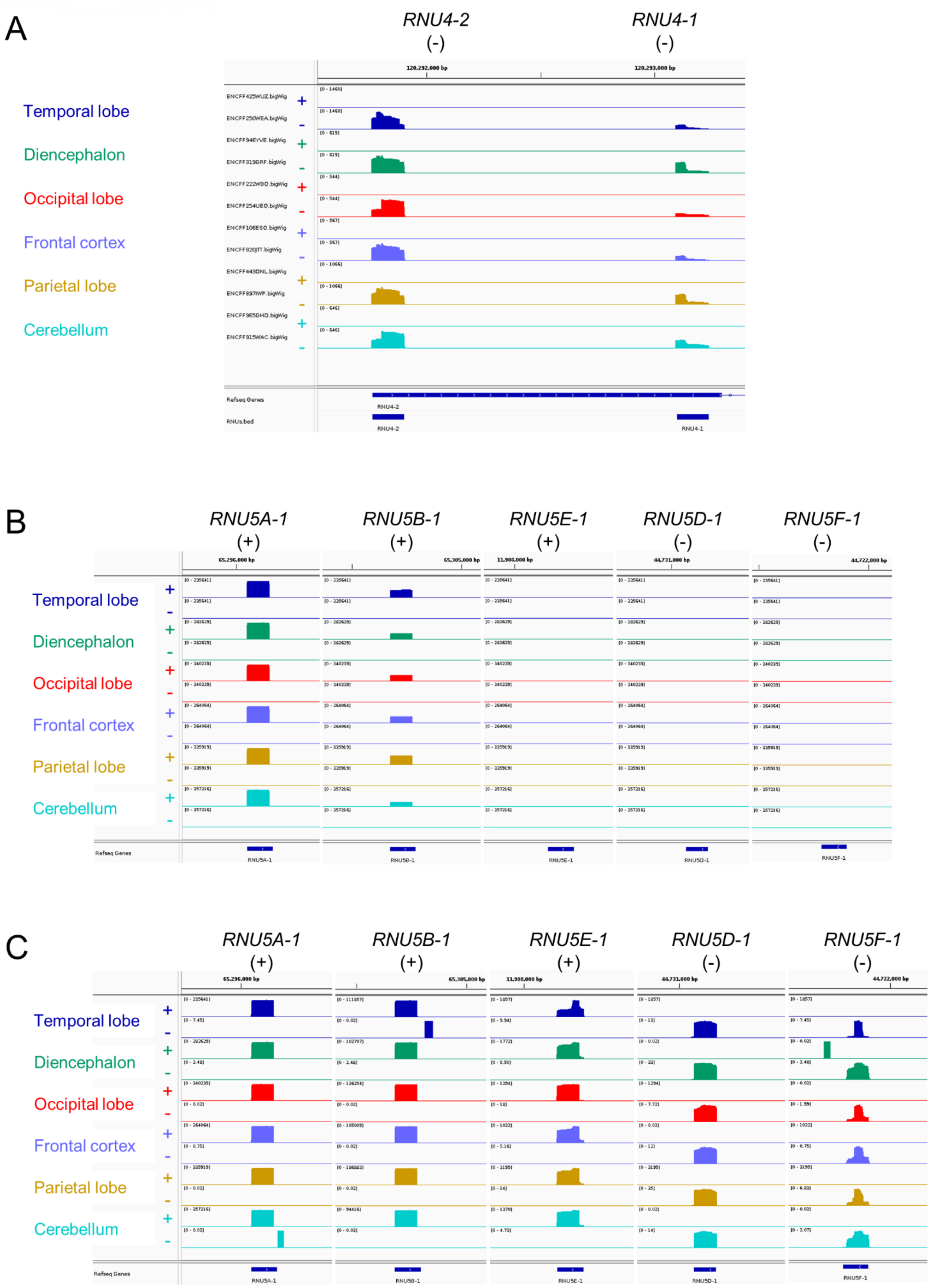


**Supplementary Figure 3: Expression of the different genes coding for U4 and U5 genes in multiple brain regions. (A)** *RNU4-2* is more expressed than *RNU4-1* in the various brain regions. Minus strand tracks (-) were auto-scaled and each of their maximum was set to the plus strand (+). **(B and C)** *RNU5A-1* and *RNU5B-*

*1* are both highly expressed in the brain, while *RNU5E-1* is much less expressed. The expression of *RNU5D-1* and *RNU5F-1* in the brain is negligible. **(B)** Plus strand track maximum at *RNU5A*-1 from each tissue was set to the minus strand and kept for all genes. **(C)** Auto-scale was allowed for each tissue and gene. Small-RNA data was generated by the ENCODE Consortium for different human embryonic brain regions[38]: diencephalon (GEO:GSE78292), temporal lobe (GEO:GSE78303), occipital lobe (GEO:GSE78298), frontal cortex (GEO:GSE78293), parietal lobe (GEO:GSE78299), cerebellum (GEO:GSE78291). Tracks show unique read signals for plus and minus strand from the default anisogenic replicate.

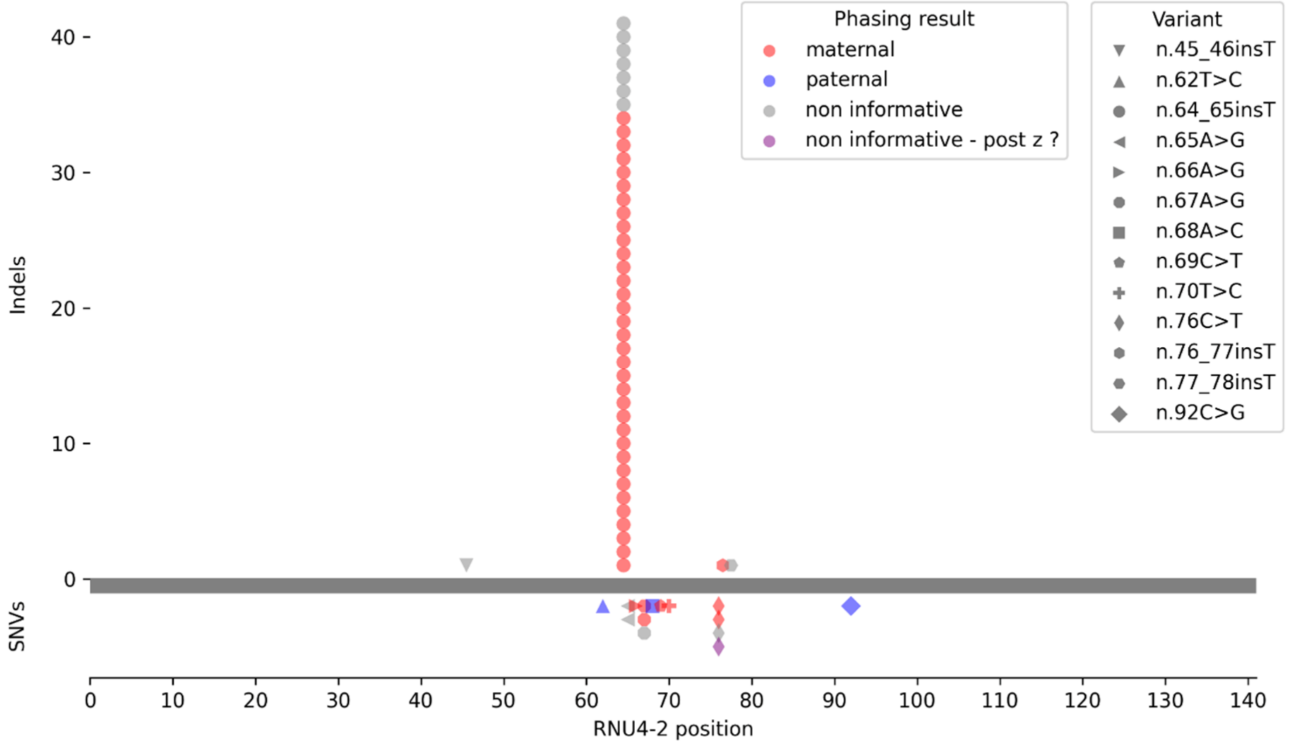


**Supplementary Figure 4: Parental haplotype of origin of *de novo RNU4-2* variants.** Haplotype of origin of *de novo RNU4-2* single nucleotide variants (SNVs) and indels detected by trio genome sequencing could be assessed using nearby inherited SNVs (*n*=59). *De novo* indels, and notably the recurrent n.64_65insT, display complete maternal bias, while SNVs were occasionally observed to happen on the paternal haplotype. Notably, a substantial proportion of *de novo* variants were successfully phased (45 out of 59, 76%), a stark contrast to the genome-wide phaseability of *de novo* mutations by short-read genome sequencing, which is approximately 30%. One variant (n.76C>T) could not be phased but displayed a biased variant allele fraction (28%, 9/32) likely reflecting early embryonic mosaicism. Of note, this plot presents pathogenic and likely pathogenic *de novo* variants, but also two variants of unknown significance, n.45_46insT and n.92C>G.

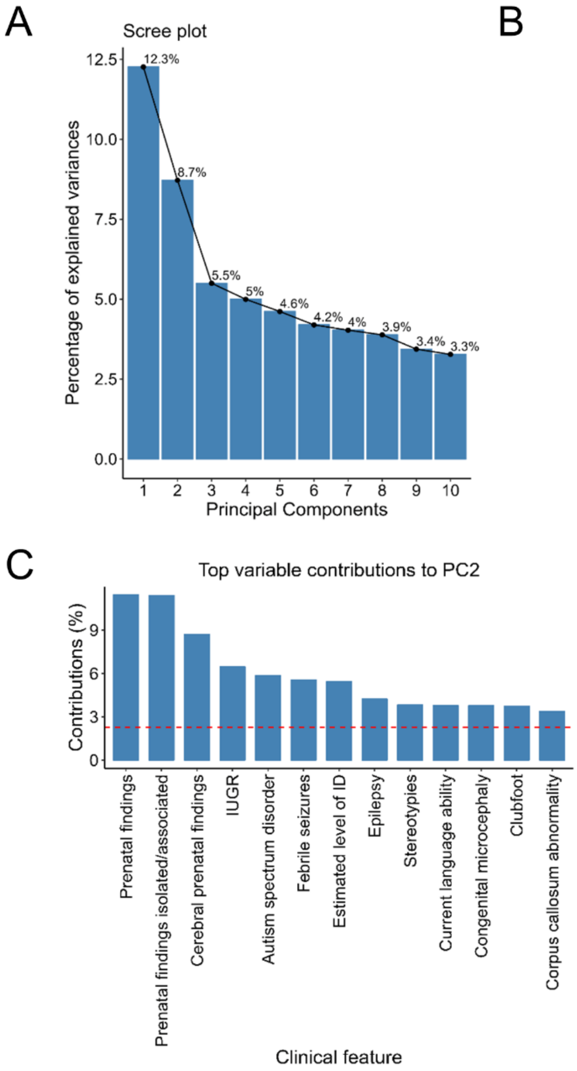


**Supplementary Figure 5: Details of the principal component analysis of clinical features associated with *RNU4-2* LP/P variants. (A)** Scree plot showing that the two first principal components explain a much larger proportion of the variance compared to the other. **(B)** Top clinical features contributing to PC1. **(C)** Top clinical features contributing to PC2. The horizontal red line represents the expected level of contribution if the contributions were uniform. Only variables with values above the red line are shown.

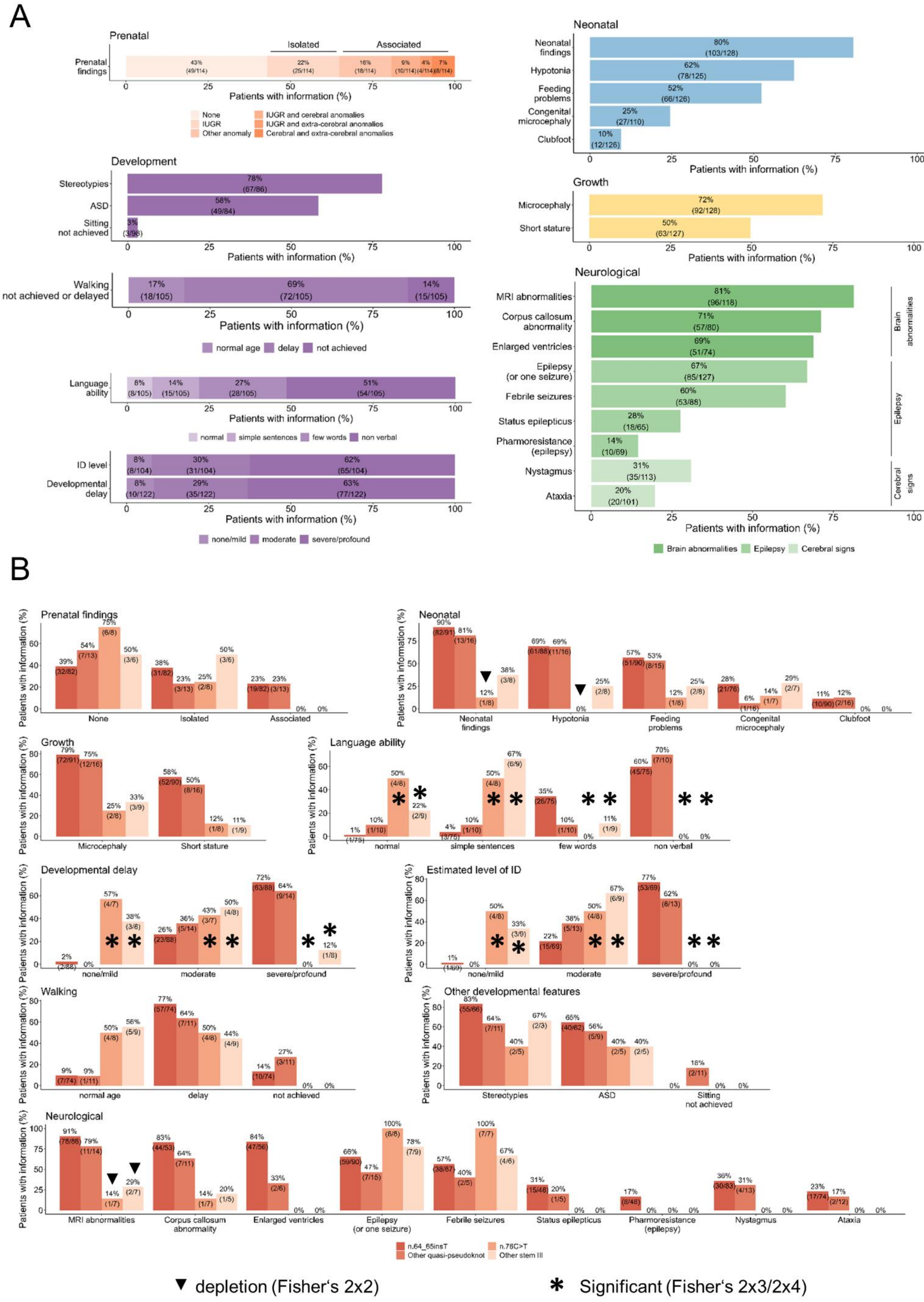

A
Prenatal
Isolated
Associated
Prenatal findings
Patients with information (%)
None
IUGR
Other anomaly
IUGR and cerebral anomalies
IUGR and extra-cerebral anomalies
Cerebral and extra-cerebral anomalies
Development
Stereotypies
ASD
Sitting not achieved
Walking not achieved or delayed
normal age
delay
not achieved
Language ability
normal
simple sentences
few words
non verbal
ID level
Developmental delay
none/mild
moderate
severe/profound
Neonatal
Neonatal findings
Hypotonia
Feeding problems
Congenital microcephaly
Clubfoot
Growth
Microcephaly
Short stature
Neurological
MRI abnormalities
Corpus callosum abnormality
Enlarged ventricles
Epilepsy (or one seizure)
Febrile seizures
Status epilepticus
Pharmoresistance (epilepsy)
Nystagmus
Ataxia
Brain abnormalities
Epilepsy
Cerebral signs
B
Prenatal findings
Neonatal
Growth
Language ability
Developmental delay
Estimated level of ID
Walking
Other developmental features
Neurological
Patients with information (%)
n.64_65insT
Other quasi-pseudoknot
n.76C>T
Other stem III
▼ depletion (Fisher's 2x2)
* Significant (Fisher's 2x3/2x4)

**Supplementary Figure 6: Overview of the clinical characteristics of patients with *RNU4-2* LP/P variants. (A)** Aggregated clinical features of the whole cohort. **(B)** Comparison of phenotypes related to *RNU4-2* variants in the T loop and stem III domains. Depletion (triangles; 2×2 contingency tables) and significant difference (asterisks; 2×3 or 2×4 contingency tables) refer to the Fisher's tests (two-sided) adjusted for multiple comparisons using Bonferroni correction presented in Table 1. Details of statistical tests appear in Supplementary Table 7.

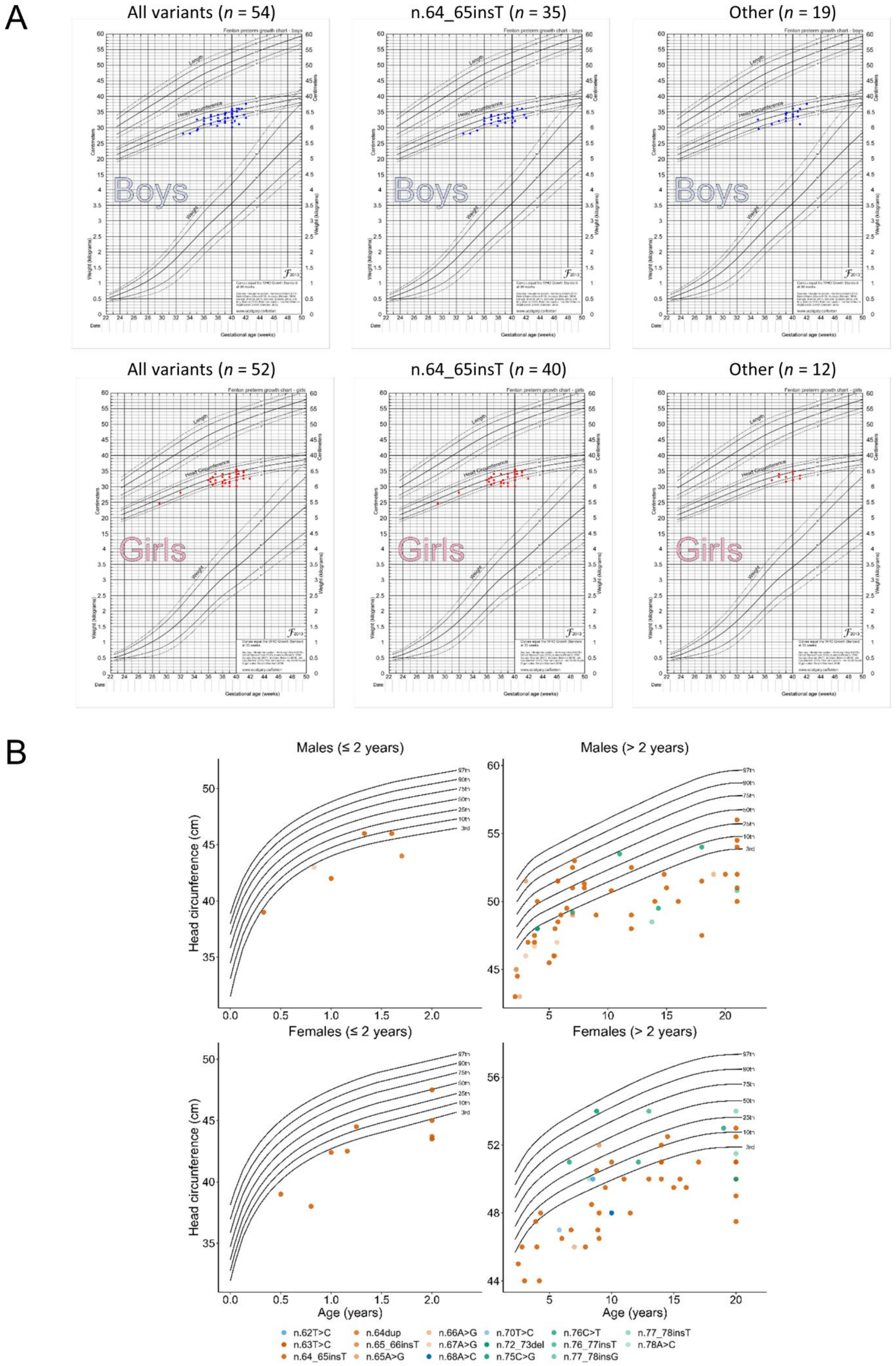


**Supplementary Figure 7: Head circumference data of patients with *RNU4-2* LP/P variants. (A)** Preterm growth charts[32] showing head circumference measurements at birth according to the gestational age. Separate graphs are shown for males (top; blue dots) and females (bottom; red dots). Graphs are shown for all pathogenic and likely pathogenic *RNU4-2* variants (left), for the recurrent n.64_65insT (middle) and for the remaining variants (right). **(B)** Head circumference measurements according to the age for patients with two years-old or less (left) or older than two years. Separate graphs are shown for males (top) and females (bottom). Reference chart data points from birth to 21 years and up (males) or to 20 years and up (females) were obtained from Rollins et al. 2010[33]. Male patients older than 21 years were plotted at age 21 and female patients older than 20 years were plotted at age 20.

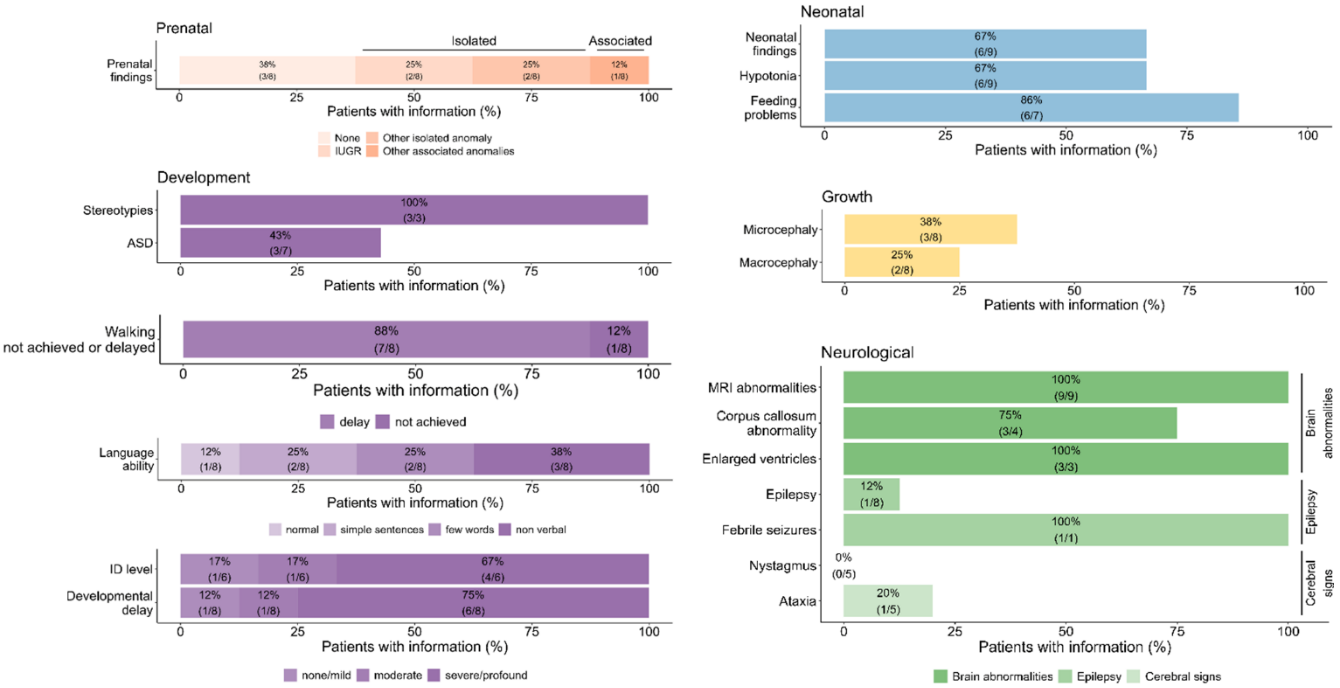


**Supplementary Figure 8: Overview of the clinical features of patients with *RNU5B-1* LP/P variants.**

A

| Input Sample | B cells naive | B cells memory | Plasma cells | T cells CD8 | T cells CD4 naive | T cells CD4 memory resting | T cells CD4 memory activated | T cells follicular helper | T cells regulatory (Tregs) | T cells gamma delta | NK cells resting | NK cells activated | Monocytes | Macrophages M0 | Macrophages M1 | Macrophages M2 | Dendritic cells resting | Dendritic cells activated | Mast cells resting | Mast cells activated | Eosinophils | Neutrophils | P-value | Correlation |
|---|---|---|---|---|---|---|---|---|---|---|---|---|---|---|---|---|---|---|---|---|---|---|---|---|
| CTR1 | 0.091 | 0 | 0 | 0.095 | 0 | 0 | [illegible] | 0 | 0.005 | 0 | 0.035 | 0 | 0.002 | 0.022 | 0 | 0.007 | 0 | 0.045 | 0 | 0.007 | 0 | 0.000 | 0.000 | 0.699 |
| CTR10 | 0.099 | 0 | 0 | 0.124 | 0 | 0 | [illegible] | 0.014 | 0.097 | 0 | 0.04 | 0 | 0.003 | 0.019 | 0 | 0.001 | 0 | 0.026 | 0 | 0 | 0 | 0.000 | 0.000 | 0.665 |
| CTR11 | 0.07 | 0 | 0 | 0.066 | 0 | 0 | [illegible] | 0 | 0.026 | 0 | 0.029 | 0 | 0.002 | 0.019 | 0 | 0.007 | 0 | 0.037 | 0 | 0.003 | 0 | 0.000 | 0.000 | 0.720 |
| CTR12 | 0.105 | 0 | 0 | 0.06 | 0.07 | 0 | [illegible] | 0.01 | 0.091 | 0 | 0.072 | 0 | 0 | 0.045 | 0 | 0.002 | 0 | 0.044 | 0 | 0.005 | 0 | 0.000 | 0.000 | 0.632 |
| CTR13 | 0.073 | 0 | 0.007 | 0.143 | 0.013 | 0 | [illegible] | 0.012 | 0.167 | 0 | 0.043 | 0 | 0.005 | 0.021 | 0 | 0 | 0 | 0.006 | 0 | 0 | 0 | 0.000 | 0.000 | 0.640 |
| CTR14 | 0.073 | 0 | 0 | 0.094 | 0 | 0 | [illegible] | 0 | 0.072 | 0 | 0.041 | 0 | 0.005 | 0.018 | 0.001 | 0.008 | 0 | 0.031 | 0 | 0.002 | 0 | 0.000 | 0.000 | 0.668 |
| CTR15 | 0.078 | 0 | 0 | 0.166 | 0 | 0 | [illegible] | 0.015 | 0.109 | 0 | 0.071 | 0 | 0.011 | 0.03 | 0 | 0.003 | 0 | 0.01 | 0.04 | 0 | 0 | 0.002 | 0.000 | 0.696 |
| CTR16 | 0.077 | 0 | 0 | 0.052 | 0.01 | 0 | [illegible] | 0.02 | 0.081 | 0 | 0.041 | 0 | 0 | 0.044 | 0.001 | 0 | 0 | 0.062 | 0 | 0.035 | 0 | 0.000 | 0.000 | 0.703 |
| CTR17 | 0.093 | 0 | 0.006 | 0.099 | 0.011 | 0 | [illegible] | 0.012 | 0.142 | 0 | 0.066 | 0 | 0.004 | 0.015 | 0 | 0 | 0 | 0.021 | 0 | 0 | 0 | 0.000 | 0.000 | 0.755 |
| CTR18 | 0.08 | 0 | 0.007 | 0.061 | 0.045 | 0 | [illegible] | 0.006 | 0.152 | 0 | 0.097 | 0 | 0.002 | 0.023 | 0 | 0.001 | 0 | 0.016 | 0 | 0 | 0 | 0.000 | 0.000 | 0.572 |
| CTR19 | 0.068 | 0 | 0.007 | 0.136 | 0.004 | 0 | [illegible] | 0.005 | 0.115 | 0 | 0.051 | 0 | 0.005 | 0.02 | 0 | 0.001 | 0 | 0.014 | 0 | 0 | 0 | 0.000 | 0.000 | 0.721 |
| CTR2 | 0.07 | 0 | 0 | 0.059 | 0.008 | 0 | [illegible] | 0 | 0 | 0 | 0.072 | 0 | 0 | 0.032 | 0.002 | 0.003 | 0 | 0.042 | 0 | 0 | 0 | 0.000 | 0.000 | 0.764 |
| CTR20 | 0.121 | 0 | 0 | 0.126 | 0 | 0 | [illegible] | 0.005 | 0.081 | 0 | 0.034 | 0 | 0.006 | 0.012 | 0.001 | 0.003 | 0 | 0.023 | 0 | 0 | 0 | 0.000 | 0.000 | 0.656 |
| CTR21 | 0.07 | 0 | 0 | 0.049 | 0 | 0 | [illegible] | 0 | 0.064 | 0 | 0.026 | 0 | 0.001 | 0.021 | 0 | 0.002 | 0 | 0.039 | 0 | 0.008 | 0 | 0.000 | 0.000 | 0.736 |
| CTR3 | 0.102 | 0 | 0.006 | 0.183 | 0.006 | 0 | [illegible] | 0.013 | 0.17 | 0 | 0.066 | 0 | 0.009 | 0.028 | 0 | 0 | 0 | 0.015 | 0 | 0 | 0 | 0.000 | 0.000 | 0.712 |
| CTR4 | 0.094 | 0 | 0.007 | 0.145 | 0.043 | 0 | [illegible] | 0.007 | 0.179 | 0 | 0.063 | 0 | 0.007 | 0.015 | 0 | 0 | 0 | 0.009 | 0 | 0 | 0 | 0.000 | 0.000 | 0.713 |
| CTR5 | 0.059 | 0 | 0 | 0.102 | 0 | 0 | [illegible] | 0 | 0.064 | 0 | 0.036 | 0 | 0.002 | 0.071 | 0 | 0 | 0 | 0.036 | 0 | 0.01 | 0 | 0.000 | 0.000 | 0.688 |
| CTR6 | 0.075 | 0 | 0.001 | 0.168 | 0.026 | 0 | [illegible] | 0.046 | 0.125 | 0 | 0.094 | 0 | 0.007 | 0.04 | 0 | 0.003 | 0 | 0.01 | 0 | 0.001 | 0 | 0.000 | 0.000 | 0.740 |
| CTR7 | 0.106 | 0 | 0.002 | 0.13 | 0 | 0 | [illegible] | 0.05 | 0.166 | 0 | 0.083 | 0 | 0 | 0.116 | 0.002 | 0.021 | 0.002 | 0.018 | 0.013 | 0 | 0 | 0.004 | 0.000 | 0.660 |
| CTR8 | 0.084 | 0 | 0.003 | 0.089 | 0.023 | 0 | [illegible] | 0.028 | 0.211 | 0 | 0.058 | 0 | 0 | 0.088 | 0 | 0 | 0.011 | 0.025 | 0.003 | 0 | 0 | 0.000 | 0.000 | 0.659 |
| CTR9 | 0.116 | 0 | 0 | 0.097 | 0.027 | 0 | [illegible] | 0.019 | 0.16 | 0 | 0.057 | 0 | 0.004 | 0.083 | 0 | 0.004 | 0 | 0.038 | 0.001 | 0 | 0 | 0.000 | 0.000 | 0.613 |
| RNU1 | 0.048 | 0 | 0 | 0.02 | 0 | 0.041 | 0.061 | 0.007 | 0.008 | 0 | 0.022 | 0 | 0 | [illegible] | 0.003 | 0.198 | 0.009 | 0.075 | 0 | 0.087 | 0 | 0.002 | 0.000 | 0.788 |
| RNU10 | 0.075 | 0 | 0 | 0.095 | 0.017 | 0 | [illegible] | 0.003 | 0.082 | 0 | 0.037 | 0 | 0.004 | 0.077 | 0 | 0.001 | 0 | 0.065 | 0 | 0.006 | 0 | 0.000 | 0.000 | 0.652 |
| RNU11 | 0.073 | 0 | 0 | 0.095 | 0.022 | 0 | [illegible] | 0 | 0.038 | 0 | 0.077 | 0 | 0 | 0.07 | 0.003 | 0.025 | 0 | 0.055 | 0.018 | 0 | 0 | 0.005 | 0.000 | 0.707 |
| RNU12 | 0.085 | 0 | 0 | 0.119 | 0.002 | 0 | [illegible] | 0.002 | 0.081 | 0 | 0.058 | 0 | 0.002 | 0.06 | 0 | 0.007 | 0 | 0.054 | 0 | 0.007 | 0 | 0.000 | 0.000 | 0.687 |
| RNU13 | 0.067 | 0 | 0 | 0.09 | 0 | 0 | [illegible] | 0 | 0.042 | 0 | 0.048 | 0 | 0.002 | 0.026 | 0 | 0 | 0 | 0.024 | 0 | 0 | 0 | 0.000 | 0.000 | 0.709 |
| RNU14 | 0.097 | 0 | 0 | 0.103 | 0.051 | 0 | [illegible] | 0 | 0.073 | 0 | 0.069 | 0 | 0.003 | 0.088 | 0.001 | 0.006 | 0 | 0.025 | 0 | 0 | 0 | 0.000 | 0.000 | 0.666 |
| RNU15 | 0.062 | 0 | 0.006 | 0.101 | 0.023 | 0 | [illegible] | 0.014 | 0.147 | 0 | 0.039 | 0 | 0.004 | 0.002 | 0 | 0 | 0 | 0.003 | 0.001 | 0 | 0 | 0.000 | 0.000 | 0.736 |
| RNU16 | 0.082 | 0 | 0 | 0.069 | 0.05 | 0 | [illegible] | 0.008 | 0.146 | 0 | 0.062 | 0 | 0 | 0.025 | 0 | 0 | 0 | 0.027 | 0.004 | 0 | 0 | 0.000 | 0.000 | 0.682 |
| RNU17 | 0.078 | 0 | 0 | 0.061 | 0.006 | 0 | [illegible] | 0.008 | 0.074 | 0 | 0.046 | 0 | 0.001 | 0.012 | 0 | 0 | 0 | 0.027 | 0 | 0.006 | 0 | 0.000 | 0.000 | 0.733 |
| RNU2 | 0.105 | 0 | 0 | 0.078 | 0 | 0 | [illegible] | 0 | 0.11 | 0 | 0.057 | 0 | 0.004 | 0.013 | 0 | 0.008 | 0 | 0.023 | 0 | 0 | 0 | 0.000 | 0.000 | 0.633 |
| RNU3 | 0.053 | 0 | 0.01 | 0.077 | 0.063 | 0 | [illegible] | 0.055 | 0.248 | 0 | 0.084 | 0 | 0 | 0.032 | 0 | 0 | 0 | 0.004 | 0.003 | 0 | 0 | 0.000 | 0.000 | 0.696 |
| RNU4 | 0.076 | 0 | 0 | 0.066 | 0 | 0 | [illegible] | 0 | 0.082 | 0 | 0.048 | 0 | 0.004 | 0.041 | 0 | 0 | 0 | 0.048 | 0 | 0.009 | 0 | 0.000 | 0.000 | 0.686 |
| RNU5 | 0.067 | 0 | 0.002 | 0.127 | 0.038 | 0 | [illegible] | 0.049 | 0.193 | 0 | 0.064 | 0 | 0.005 | 0.076 | 0 | 0 | 0 | 0.005 | 0 | 0 | 0 | 0.000 | 0.000 | 0.659 |
| RNU6 | 0.042 | 0 | 0 | 0.061 | 0 | 0 | [illegible] | 0 | 0 | 0 | 0.028 | 0 | 0 | 0.16 | 0.035 | 0.019 | 0 | 0.069 | 0 | 0.031 | 0 | 0.000 | 0.000 | 0.618 |
| RNU7 | 0.073 | 0 | 0 | 0.071 | 0.02 | 0 | [illegible] | 0 | 0.037 | 0 | 0.053 | 0 | 0 | 0.204 | 0.006 | 0.008 | 0 | 0.029 | 0 | 0.071 | 0 | 0.000 | 0.000 | 0.631 |
| RNU8 | 0.085 | 0 | 0 | 0.109 | 0 | 0 | [illegible] | 0 | 0.043 | 0 | 0.033 | 0 | 0.003 | 0.077 | 0 | 0.004 | 0 | 0.046 | 0 | 0.007 | 0 | 0.000 | 0.000 | 0.682 |
| RNU9 | 0.075 | 0 | 0.003 | 0.123 | 0.006 | 0 | [illegible] | 0.009 | 0.095 | 0 | 0.055 | 0 | 0.003 | 0.086 | 0 | 0 | 0 | 0.042 | 0 | 0.002 | 0 | 0.000 | 0.000 | 0.685 |

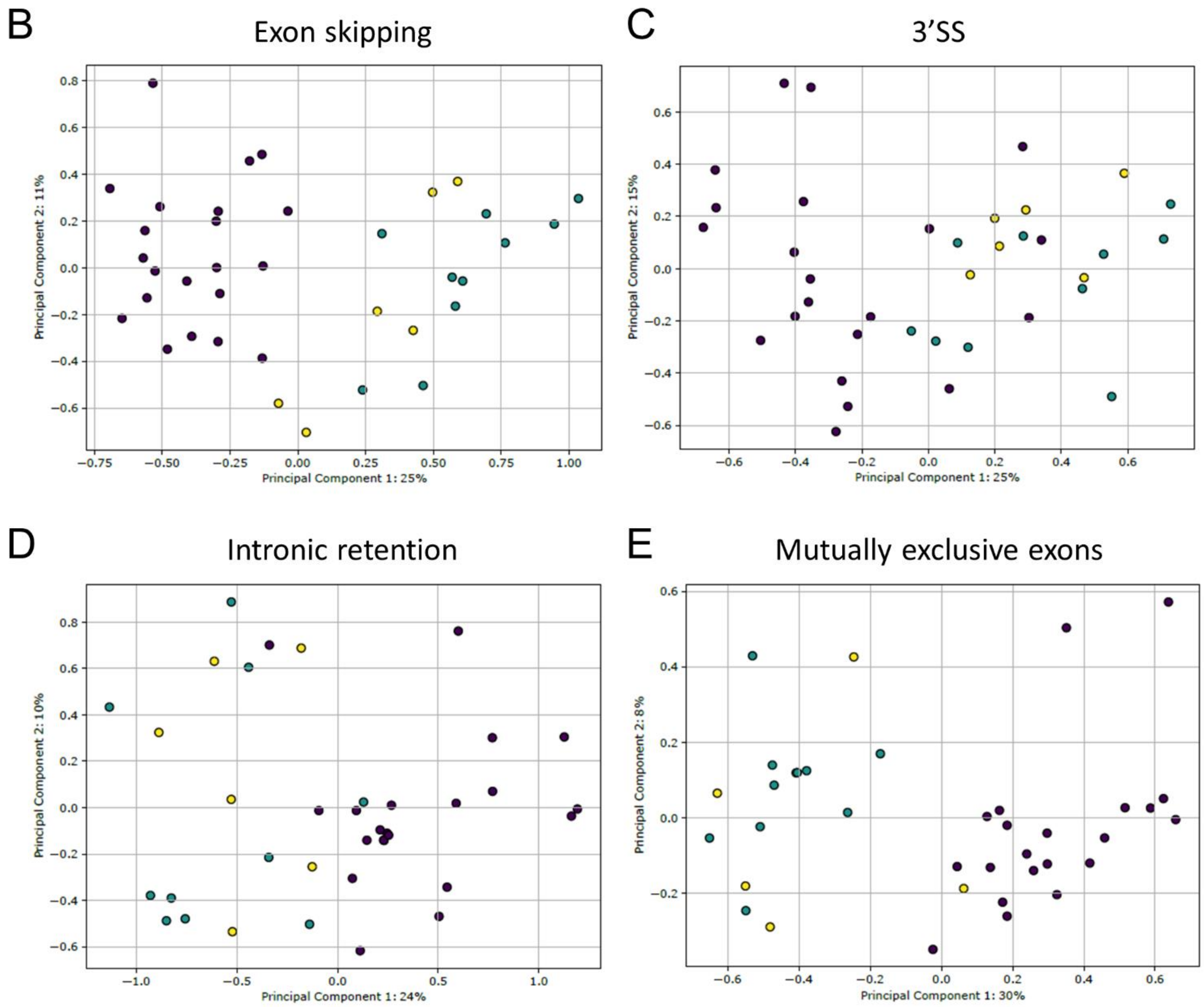

**Supplementary Figure 9: CIBERSORTx quality control and PCA of rMATS output other than alternative 5' splice sites (5'SS). (A)** CIBERSORTx imputation of blood cells abundance in short-term lymphocyte cultures. Imputations were performed using LM22 signature matrix. One patient sample was removed from further analysis based on a low proportion of activated T CD4+ cells compared to other *RNU4-2* variants and controls. The sample name is anonymized and cannot be used to identify the corresponding individual. **(B-E)** PCA of rMATS output other than 5'SS. PCA were performed using PSI values of significant calls (FDR<0.1) with a |deltaPSI| > 0.05 for exon skipping (B, *n*=119), alternative 3' splice sites (C, *n*=32, 3'SS), intronic retention (D, *n*=81), mutually exclusive exons (E, *n*=102). Purple: controls, Green: *RNU4-2* n.64_65insT, Yellow: other variants.

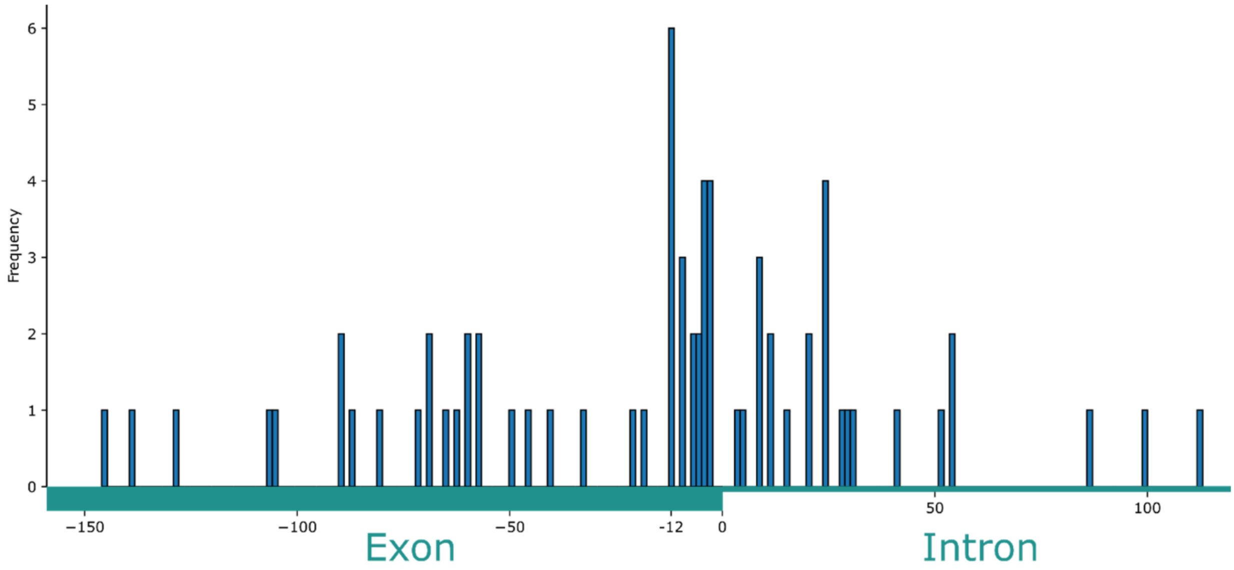


**Supplementary Figure 10: Distance between the end of the exon and the 5'SS.**

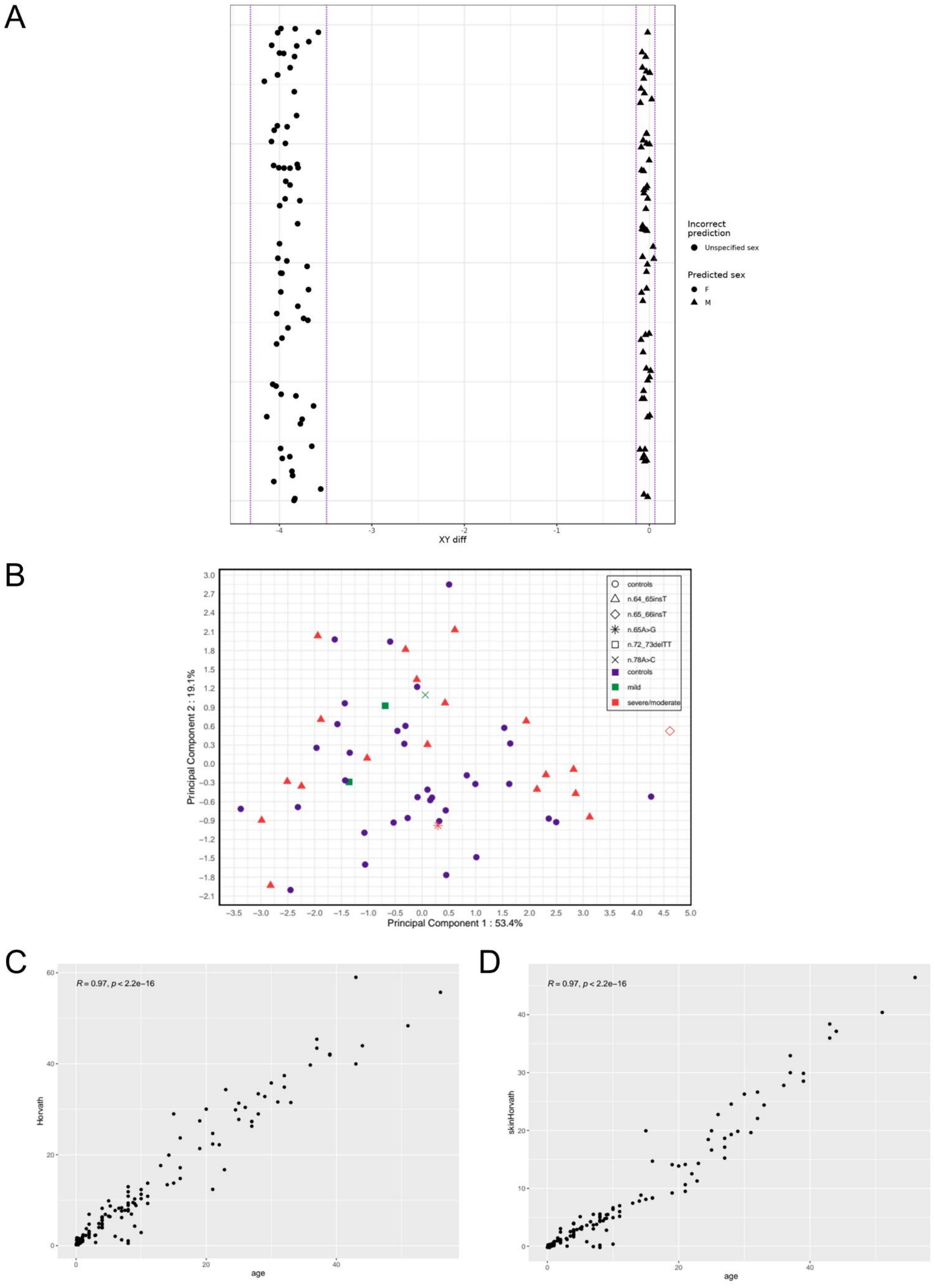


**Supplementary Figure 11: Age, sex and cell composition controls performed for DNA methylation data.** **(A)** Methylation levels show perfect concordance between reported and predicted sex. Plot of the difference between median chromosome Y and chromosome X probe intensities ("XY diff") as obtained by meffil standard QC. Cutoff for sex detection was XY diff = -2. Mismatched samples are shown in red. The dashed lines represent 3 standard deviations (SD) from the mean xy difference. Samples in this interval are considered outliers. **(B)** Patients with *RNU4-2* variants display similar estimated blood cell count distributions as controls on methylation data, with perfect overlap on first principal components. The predicted cellular composition consists of six elements (B cells, CD4T+ cells, CD8T+ cells, NK cells, Granulocytes and Monocytes). Each variation is represented by a distinct symbol. **(C and D)** Correlation between actual age at blood sample and predicted age with Horvath clock (C) and with skinHorvath clock (D). Pearson correlation was used.

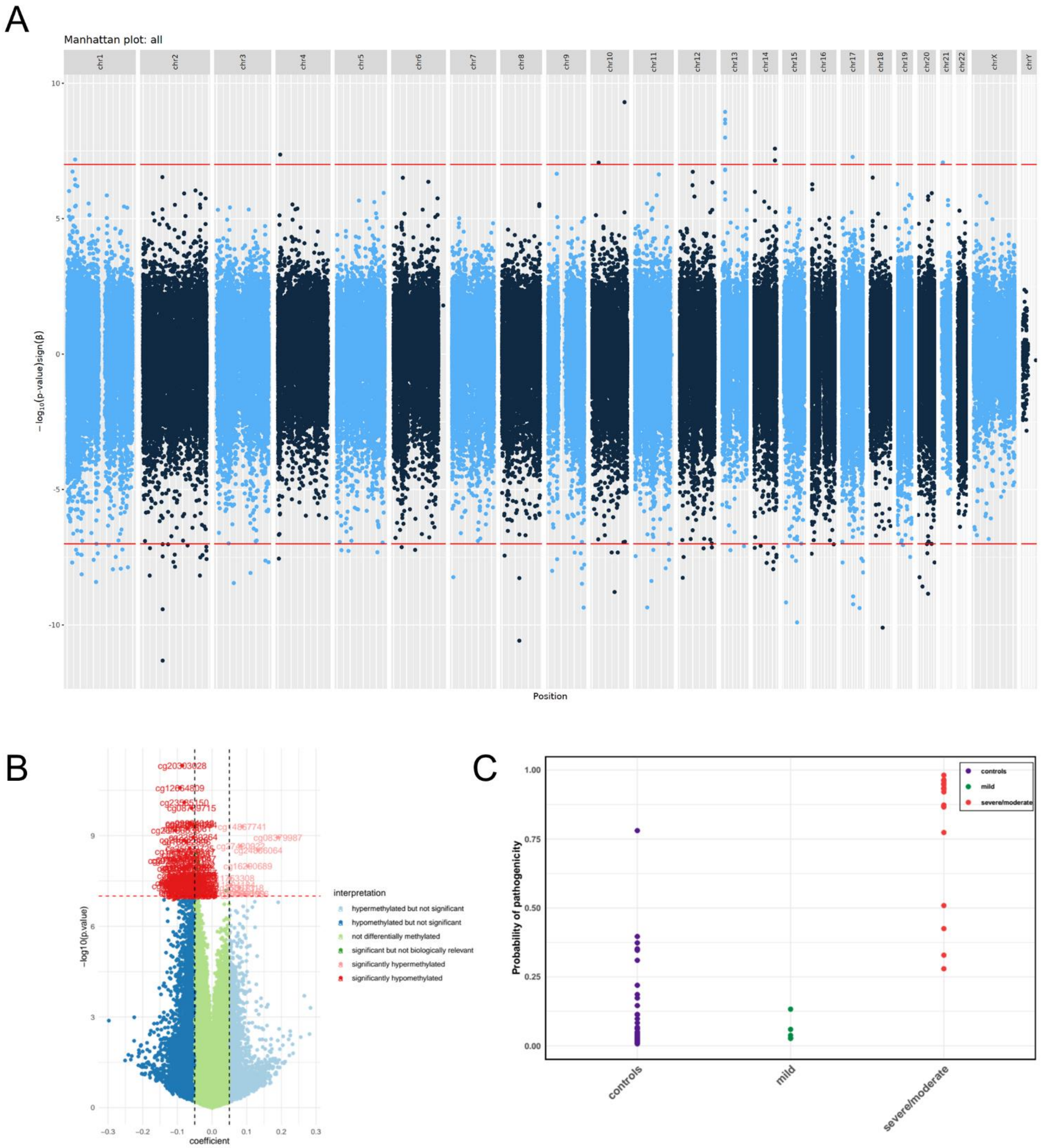


**Supplementary Figure 12: Epigenetic study. (A and B)** Epigenome-wide association study (EWAS) Manhattan (A) and volcano plots (B). Probes used for the signature are represented in pink and red on the volcano plot, indicating hypo and hypermethylated probes, respectively with uncorrected *p*-value < $10^{-7}$ and |Δβ| > 5%. **(C)** Classification of samples by SVM based on DNAm signature after 5-fold cross-validation. Phenotypes are displayed on the x-axis, while the y-axis displays the probability of pathogenicity.
